# Artificial Intelligence-Enhanced Comprehensive Assessment of the Aortic Valve Stenosis Continuum in Echocardiography

**DOI:** 10.1101/2024.07.08.24310123

**Authors:** Jiesuck Park, Jiyeon Kim, Jaeik Jeon, Yeonyee E. Yoon, Yeonggul Jang, Hyunseok Jeong, Youngtaek Hong, Seung-Ah Lee, Hong-Mi Choi, In-Chang Hwang, Goo-Yeong Cho, Hyuk-Jae Chang

## Abstract

**Background:** Transthoracic echocardiography (TTE) is the primary modality for diagnosing aortic stenosis (AS), yet it requires skilled operators and can be resource-intensive. We developed and validated an artificial intelligence (AI)-based system for evaluating AS that is effective in both resource-limited and advanced settings.

**Methods:** We created a dual-pathway AI system for AS evaluation using a nationwide echocardiographic dataset (developmental dataset, n=8,427): 1) a deep learning (DL)-based AS continuum assessment algorithm using limited 2D TTE videos, and 2) automating conventional AS evaluation. We performed internal (internal test dataset [ITDS], n=841) and external validation (distinct hospital dataset [DHDS], n=1,696; temporally distinct dataset [TDDS], n=772) for diagnostic value across various stages of AS and prognostic value for composite endpoints (cardiovascular death, heart failure, and aortic valve replacement)

**Findings:** The DL index for the AS continuum (DLi-ASc, range 0-100) increases with worsening AS severity and demonstrated excellent discrimination for any AS (AUC 0.91– 0.99), significant AS (0.95–0.98), and severe AS (0.97–0.99). DLi-ASc was independent predictor for composite endpoint (adjusted hazard ratios 2.19, 1.64, and 1.61 per 10-point increase in ITDS, DHDS, and TDDS, respectively). Automatic measurement of conventional AS parameters demonstrated excellent correlation with manual measurement, resulting in high accuracy for AS staging (98.2% for ITDS, 82.1% for DHDS, and 96.8% for TDDS) and comparable prognostic value to manually-derived parameters.

**Interpretation:** The AI-based system provides accurate and prognostically valuable AS assessment, suitable for various clinical settings. Further validation studies are planned to confirm its effectiveness across diverse environments.

**Research in Context:** *Evidence before this study:* We screened all English-based research articles in PubMed up to December 2023 using the keywords “artificial intelligence," "echocardiography," “aortic stenosis,” and "aortic valve stenosis." While some studies have used artificial intelligence (AI) to evaluate aortic stenosis (AS) in echocardiography, these efforts were typically focused on either predicting significant AS or automating conventional measurements, not both. For instance, Wesler BS et al. trained a deep learning model on 338 patients and validated it with 119 patients, achieving an area under the receiver operating characteristic curve (AUC) of 0.86 for distinguishing significant AS from non-significant AS. In a larger-scale study, Holste G. et al. trained a deep learning model on 5,257 studies and validated it using two external datasets (4,226 and 3,072 studies), achieving high accuracy in detecting severe AS (AUC: 0.942–0.952). However, both models were limited to the parasternal long-axis view and did not provide conventional quantitative analysis. In contrast, Krishna H. et al. automated conventional AS evaluation, demonstrating that AI could accurately measure AS parameters like aortic valve maximal velocity, mean pressure gradient, and aortic valve area in 256 patients, comparable to human measurements, but did not perform qualitative assessment of AS. Additionally, while Strange G et al. identified AI-based AS phenotypes linked to mortality risk using data from echocardiographic reports, this approach was based on tabular data rather than direct image analysis, thus lacking the capability to assess AS severity from imaging data.

*Added value of this study:* In this study, we developed a comprehensive AI-based system to evaluate AS through a dual pathway: 1) assessing AS presence and severity by deriving a DL index for the AS continuum (DLi-ASc) from parasternal long and/or short axis videos only, and 2) automatically measuring AS parameters and providing conventional quantitative AS evaluation if additional images are available. The system was validated internally and in two independent external datasets, where DLi-ASc increased with AS severity and demonstrated excellent discrimination for any AS (AUC 0.91–0.99), significant AS (0.95–0.98), and severe AS (0.97–0.99). Additionally, DLi-ASc independently predicted adverse cardiovascular events. The automatic measurement of conventional AS parameters showed a strong correlation with manual measurement, resulting in high accuracy for AS staging (98.2% for internal test set, 81.0%, and 96.8% for external test sets) and offered prognostic value comparable to manually-derived parameters.

*Implications of all the available evidence:* AI-enhanced echocardiographic evaluation of AS allows for accurate diagnosis of significant AS and prediction of severity using only parasternal long or short axis views, typically obtained in the first step of echocardiographic evaluation. This capability can enhance AS assessment in resource-limited settings and provide novices with guidance on when quantitative analysis is necessary. If additional views are appropriately acquired, the system automatically analyses them, potentially enabling conventional quantitative evaluation, thereby saving time and effort while ensuring accurate assessment. However, further comparative prospective studies are necessary to assess whether this AI-based approach ensure these efficiencies without inadvertently increasing diagnostic errors or adverse cardiac outcomes compared to conventional, manual AS evaluation.

## 1. Introduction

With the global increase in aging populations, the prevalence of degenerative disease like aortic stenosis (AS) is rising, necessitating timely detection and management to prevent severe outcomes.^1,2^ Transthoracic echocardiography (TTE) is the primary imaging modality for assessing AS, offering generally reliable results. However, accurate AS staging requires advanced equipment and expertise, particularly for assessment with multiple measurements and Doppler analyses. In nonspecialized settings, such as those limited to point-of-care ultrasound (POCUS), even Doppler acquisition is not feasible, highlighting the potential of AI to assess AS using limited images. Even in tertiary care centres, the process remains labour-intensive, underscoring the need for automated solutions that streamline AS evaluation by handling measurements and simplifying the interpretation process.

Previous studies have typically focused on classifying significant or severe AS using limited TTE image-most commonly a single parasternal long (PLAX) axis view^3,4^ – or have proposed AI-based automation of conventional quantitative analyses, assuming access to advanced imaging setups.^5^ In contrast, we developed a comprehensive artificial intelligence (AI)-based system designed for applicability in both resource-limited and advanced settings. Leveraging deep learning (DL), our system assesses AS using only a limited 2-dimensional (2D) TTE videos – PLAX and/or parasternal short-axis (PSAX) views. Unlike previous models that focus solely on classification of significant or severe AS our system is designed to reflect the full severity continuum of AS. Simultaneously, it also automates the measurements of a broad range of structural and hemodynamic parameters, enabling the conventional calculation of the aortic valve area (AVA) for a quantitative assessment of AS. This paper details the development of our AI-based system and demonstrates its diagnostic and prognostic capabilities in AS assessment.

## 2. Methods

### 2.1. Study population and data sources

The AI-based frameworks utilized in this study were developed and validated using the Open AI Dataset Project (AI-Hub) dataset, an initiative supported by the South Korean government’s Ministry of Science and ICT.^6^ This dataset consists of 30,000 echocardiographic examinations retrospectively collected from five tertiary hospitals between 2012 and 2021, covering a wide range of cardiovascular diseases (***Supplemental Methods 1***). The AI-based frameworks introduced here were all developed using data extracted from the AI-Hub dataset.^7–9^ To develop the DL-based AS continuum assessment algorithm, a key focus of this study, we assembled the Development Dataset (DDS) by deliberately excluding Severance Hospital data among five hospitals. Instead, data from Severance Hospital were used exclusively for external validation (Distinct Hospital Dataset, DHDS). Further external validation was conducted using data collected from Seoul National University Bundang Hospital in 2022 (Temporally Distinct Dataset, TDDS). Detailed methodologies for data utilization in developing and validating the AI-based system are in Supplemental Methods 1. As a result, the DDS comprised TTE images from 8,427 patients, while the DHDS included 1,696 patients, and the TDDS included 772 patients. The study followed the Declaration of Helsinki (as revised in 2013). The institutional review board of each hospital approved this study and waived the requirement for informed consent because of the retrospective and observational nature of the study design (2021-0147-003 / CNUH 2021-04-032 / HYUH 2021-03-026-003 / SCHBC 2021-03-007-001/ B-2104/677-004). Additionally, to establish and utilize the TDDS, further IRB approval was obtained from Seoul National University Bundang Hospital (approved no. B-2305-827-002). All clinical and echocardiographic data were fully anonymized before data analysis.

### 2.2. Echocardiogram acquisition and interpretation

All echocardiographic studies were conducted by trained echocardiographers or cardiologists and interpreted by board-certified cardiologists specialized in echocardiography, following recent guidelines^10,11^ as part of routine clinical care. To reflect actual clinical practice, parameters were not re-measured for the study; instead, the values from the clinical reports were used as ground truth (GT) labels. AS presence and severity were determined in the DDS using the standard clinical criteria to ensure appropriate training (***Table 1***).^10^ Cases with discordant classifications – such as those where Vmax and mPG fell into different severity categories (e.g., mild-to-moderate or moderate-to-severe AS) or low-flow low-gradient (LFLG) severe AS cases with AVA <1cm^2^ but without severe-range Vmax or mPG – were excluded from training to provide more consistent GT labelling (***Supplemental Methods 1***). These cases were later used to evaluate the performance of DLi-ASc. For the DHDS and TDDS, the prior clinician’s decision regarding AS severity in the clinical report was used as is, to better reflect actual clinical practice. Consequently, cases like LFLG severe AS were classified as severe AS based on the clinician’s judgement. These cases were subsequently utilized to validate the discriminative power of DLi-ASc.

**Table 1.**
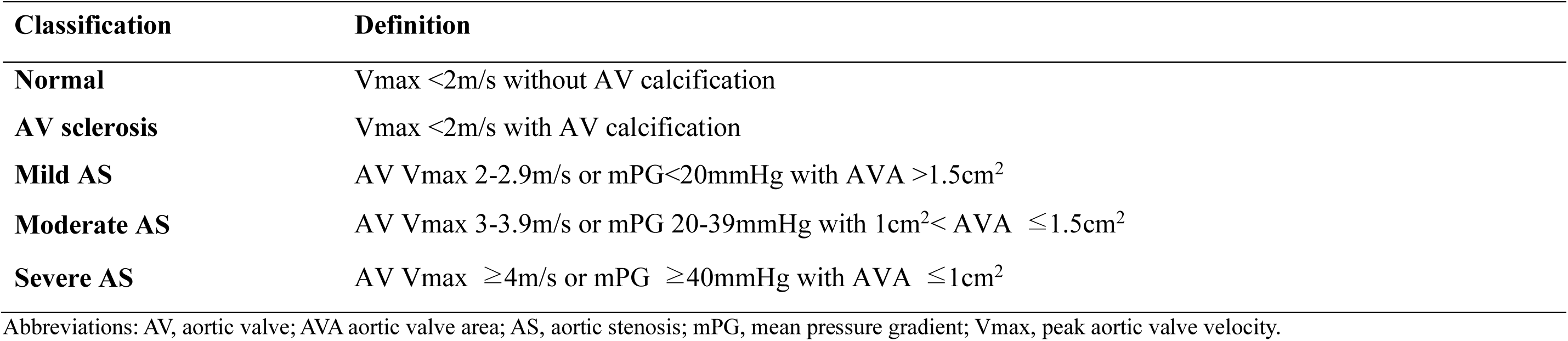
Definition of AV Staging.

| **Classification** | **Definition** |
| --- | --- |
| **Normal** | V<sub>max</sub> <2m/s without AV calcification |
| **AV sclerosis** | V<sub>max</sub> <2m/s with AV calcification |
| **Mild AS** | AV V<sub>max</sub> 2-2.9m/s or mPG<20mmHg with AVA >1.5cm<sup>2</sup> |
| **Moderate AS** | AV V<sub>max</sub> 3-3.9m/s or mPG 20-39mmHg with 1cm<sup>2</sup>< AVA ≤1.5cm<sup>2</sup> |
| **Severe AS** | AV V<sub>max</sub> ≥4m/s or mPG ≥40mmHg with AVA ≤1cm<sup>2</sup> |

Abbreviations: AV, aortic valve; AVA aortic valve area; AS, aortic stenosis; mPG, mean pressure gradient; V<sub>max</sub>, peak aortic valve velocity.

### 2.3. AI-based system

We have developed a fully automated AI-based framework that addresses AS evaluation through the dual pathway, leveraging innovative and conventional methodologies. (***Central Illustration***) The operational sequence of this system begins by automatically selecting the necessary views, including the parasternal long-axis (PLAX), parasternal short-axis (PSAX) at the aortic valve (AV) level, AV continuous wave (CW) and pulsed wave (PW) Doppler, and left ventricular outflow tract (LVOT) PW Doppler. In the DL-based AS continuum assessment pathway, the algorithm evaluates AS using only the PLAX and/or PSAX videos. Concurrently, the DL segmentation network generates masks for each view in the automated conventional AS assessment pathway. These masks facilitate the measurement of LVOT diameter from the PLAX view and analyse spectral Doppler images to ascertain key indicators such as AV peak velocity (V_max_), AV velocity time integral (VTI), AV mean pressure gradient (mPG), and LVOT VTI. Then, the system calculates AVA, enabling quantitative evaluation of AS. This AI framework represents the latest advancements in our AI-driven valve evaluation module (USfeat_valve.ai, Ontact Health, Korea), which integrates rigorously validated features such as view classification and automatic measurement capabilities.^7,9,12^ The following sections provide a detailed description of the novel component and its functionality.

#### 2.3.1. View classification

To assess AS, we improved our preexisting view classification algorithm.^7^ The algorithm could already identify the PLAX view, PSAX at the AV level, AV CW Doppler from apical views, AV PW Doppler, and LVOT PW Doppler. We augmented it to recognize the PLAX-AV zoomed views and the AV CW Doppler obtained from the right parasternal view. Detailed information about this development is in ***Supplemental Methods 2***.

#### 2.3.2. DL-based AS continuum assessment algorithm

Our objective was to develop a network that classifies AS severity in a way that reflects its continuum nature rather than just discrete categories. We used 3-dimensional (3D) convolutional neural networks (CNNs; r2plus1d18) as a backbone to separate spatial and temporal filters (***Supplemental Methods 3***).^13^ This network processes TTE videos -PLAX and/or PSAX at the AV level - to output a score predicting the AS severity, termed the DL index for the AS continuum (DLi-ASc).

To achieve accurate classification reflecting the AS continuum, we implemented bi-modal strategies: 1) continuous mapping with ordered labels and 2) multi-task learning with auxiliary tasks that predict numeric parameters indicative of the AS continuum, such as AV V_max_, mPG, and AVA. Conventional multi-class classification with cross-entropy loss was unsuitable for reflecting the AS continuum as it fails to capture the disease’s progressive nature due to equidistance between one-hot encoded severity levels. Instead, the continuous approach assigns each severity level a value between 0 and 1 (e.g., Normal: 0, Sclerosis: 0.25, Mild: 0.5, Moderate: 0.75, and Severe: 1) and trains the model by minimizing negative Bernoulli likelihood *L_Bernoulli_* While this method reflects AS progression, it primarily converts discrete labels into continuous values. To truly capture the continuum and enable nuanced transitions within and between severity levels, we incorporated three auxiliary tasks predicting TTE parameters based solely on 2D TTE videos. These tasks, predicting V_max_, mPG, and AVA, provide rich information content, allowing the network to learn anatomical features and the motion of the AV. The loss function for each auxiliary task is the mean squared error (MSE) between the predicted and actual TTE parameter values: 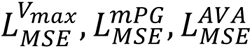. Training the network to predict continuous TTE parameters allows it to capture both discrete transitions and subtle variations within each severity category. For instance, it can distinguish between cases classified as "moderate" closer to mild AS and those nearing severe AS. The combined loss function integrates the negative Bernoulli likelihood and the MSE losses for the auxiliary tasks 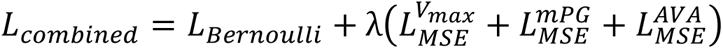, where λ is a weighting parameter balancing the contributions of the classification and regression tasks. Detailed network configurations and implementation details are in ***Supplementary Methods 3***.

Finally, to determine the patient-level DLi-ASc, if multiple PLAX or PSAX videos were available for a single patient, the DLi-ASc was extracted from each video individually. Scores from PLAX and PSAX videos were averaged separately. If only one view type (either PLAX or PSAX) was available, its average score was used directly. If both views were available, the final DLi-ASc was calculated by averaging the scores from both PLAX and PSAX views.

To develop and validate the model, we curated a Developmental Dataset (DDS) consisting of TTE data from 8,427 individuals. This dataset was divided in an 8:1:1 ratio for training, validation, and internal testing purposes. Both the training and validation sets contributed to model training, parameter tuning, and early stopping *(**Supplemental Methods 3**)*.

While the DLi-ASc was designed to capture the continuous nature of AS severity, establishing specific cutoffs is essential for clinical decision-making. To determine these cutoffs, we calculated the midpoint between the mean values of each consecutive AS severity category within the validation dataset (***Supplemental Methods 4***). This yielded the following DLi-ASc cutoffs: 24.6 for AV sclerosis, 45.4 for mild AS, 53.7 for moderate AS, and 69.7 for severe AS.

#### 2.3.3 Automated conventional AS assessment algorithm

Our AI-based system also automates the conventional method to calculate AVA and assess AS severity. Automating conventional AVA assessment in our system involves three key steps: 1) segmentation of anatomical structures and spectral Doppler envelopes, 2) uncertainty quantification to assess the confidence of the predicted segmentation masks, and 3) post-processing algorithms to extract clinical measurements from segmentation masks.

We had previously developed and validated algorithms for analysing spectral Doppler by segmenting the Doppler envelope to capture velocity profiles with essential topological features.^8,9^ This approach automatically measures AV V_max_, AV VTI, and LVOT VTI by segmenting Doppler envelopes in every analysable cycle in all provided images. In this study, to quantify AVA, we further developed a DL network based on the SegFormer transformer architecture to measure the LVOT diameter in the PLAX view.^14^ This advanced model can segment all anatomical structures visible in the PLAX view, including the left ventricle (LV), LV septum and posterior wall, left atrium, right ventricle, aorta, and even the mitral valve and AV. Detailed information is provided in ***Supplemental Methods 5*** and ***Videos S1***.

Deep segmentation networks are highly effective due to their ability to learn complex patterns and features from large datasets. However, quantifying uncertainty in their predictions is crucial because segmentation errors can impact subsequent post-processing for automatic measurement. To address this, we used predictive entropy from the segmentation network’s probability map, which combines two sources of uncertainty: lack of knowledge in DL (epistemic uncertainty) and poor data quality (aleatoric uncertainty).^15^ By evaluating the predictive entropy, we identified cases that require manual review due to poor image quality or model uncertainty, at which point the automatic measurement process was halted. Detailed methodologies are provided in ***Supplemental Methods 6*** and ***Videos S2***.

In the post-processing stage, the segmented masks were utilized to extract clinical measurements. From the predicted segmentation mask, we identified points where the mitral valve intersects with the aorta and where the septum intersects with the aorta to determine annulus points. Considering the differing opinions on the appropriate location for measuring the LVOT diameter,^16^ our algorithm was designed to measure the LVOT diameter at three different locations: at the annulus, 2.5mm, and 5mm away from the annulus towards the LV cavity. In this study, the measurements taken at the annulus were used for analysis as they showed the highest agreement with the GT. For technical details and performance information, please refer to ***Supplemental Methods 7*** and ***Video S1***.

For spectral Doppler images, AV V_max_ and VTI were derived from the segmented Doppler envelope of AV CW Doppler. This analysis included AV CW Doppler obtained from both the apical and right parasternal views, selecting the largest envelope across all cycles in all images to obtain AV V_max_ and VTI, aiming to prevent underestimation of AS jet velocity. The LVOT PW Doppler analysis also spanned all cycles, using the average value of LVOT VTI to avoid overestimating LVOT flow.^11^ These measurements were then used to calculate mPG and AVA, which were used to assess the presence and severity of AS.^10^

### 2.4 Ascertainment of clinical information and outcome definition

The clinical data were acquired through a dedicated review of electronic health records at the study institutions, including demographics (age, sex, and body mass index), comorbidities (hypertension, and diabetes), and the occurrence of clinical outcomes. The primary clinical outcome was defined as a composite endpoint of cardiovascular death, hospitalization for heart failure, and AV replacement via surgical or transcatheter approaches. Patients were censored at the occurrence of any outcome event, or at the last date of the follow-up or transferred to other institutions.

### 2.5 Validation of AI-based AS evaluation system and statistical analysis

The performance of each stage in our AI-based framework was validated using an internal test dataset (ITDS) and two external datasets (DHDS and TDDS). Additionally, we evaluated the execution time of each module, including the DLi-ASc computation, PLAX auto-measurement, and Spectral Doppler auto-measurement, across 20 repeated runs. This analysis was conducted under the following conditions: OS Windows 10, CPU Intel i7-8565U @1.80GHz, Memory 16GB, and no GPU.

The view classification algorithm, which serves as the shared initial step, was evaluated against human expert labels. Precision, recall, and F1 scores were calculated for each view, with overall accuracy determined by the ratio of correctly classified images to the total number of images. Following a manual review process, any misclassifications were corrected to ensure that subsequent analyses were performed on accurately classified views.

The performance of the DL-based AS continuum assessment algorithm was evaluated by examining the distribution of the DLi-ASc across various stages using violin plots. To verify that DLi-ASc accurately reflects the continuum of AS progression, we used Uniform Manifold Approximation and Projection (UMAP) to visualize this relationship,^17^ projecting the data into a 2D space, using 15 nearest neighbours, a minimum distance of 0.1, and the Euclidean distance. To highlight the areas with the greatest influence on the model’s prediction, we generated saliency maps using the Gradient-weighted Class Activation Mapping (Grad-CAM).^18^ We present representative samples for each severity level in both PLAX and PSAX views.

The conventional AS assessment algorithm was validated by comparing AI-derived parameters with manual measurements. Since these parameters are not typically measured in normal or AV sclerosis groups, the comparison was limited to the AS group. Moreover, as manual measurements were not always available for all AS cases, details on GT measurements availability and the success rate of automatic measurements are provided in ***Supplemental Methods 8***. The association between automated and manual measurements was assessed using the Spearman correlation analysis (r) and mean absolute error (MAE). The AS severity determined from the automatic measurements was also compared to the ground truth label made by the clinician’s prior decision.

We also evaluated the discrimination ability of the DLi-ASc and other AI-derived conventional parameters for various stages of AS, including mild or greater AS (any AS), moderate or greater AS (significant AS), and severe AS. This evaluation was conducted through receiver operating characteristic (ROC) curve analysis, from which we calculated the area under the curve (AUC).

Lastly, we assessed the prognostic capability of AI-derived parameters for composite endpoints. Specifically, we conducted a spline curve analysis for our DL index, the DLi-ASc, to visualize its predictive power. Additionally, we applied Cox regression analysis to validate the prognostic relevance of the DLi-ASc and other AI-derived AS parameters, with adjustment for clinical risk factors (age, sex, body mass index, hypertension, and diabetes).

## 3. Results

### 3.1 Baseline characteristics

Baseline clinical characteristics and the distribution of AS severity across three datasets is shown in ***Table 2***: ITDS (n=841), DHDS (n=1,696), and TDDS (n=772). ITDS and TDDS exhibited a higher prevalence of mild AS (28% and 41%, respectively), with fewer moderate and severe cases. Conversely, DHDS displayed a more balanced severity distribution (12% mild, 15% moderate, and 12% severe, respectively).

**Table 2.** Baseline Characteristics.

| Classification | Developmental Dataset (DDS) |  |  |  |  |
| --- | --- | --- | --- | --- | --- |
|  | Training<br>(n=6,743) | Validation<br>(n=843) | Internal<br>Test<br>Dataset<br>(ITDS)<br>(n=841) | Distinct Hospital<br>Dataset (DHDS)<br>(n=1,696) | Temporally<br>Distinct Dataset<br>(TDDS) (n=772) |
| **AV staging** |  |  |  |  |  |
| Normal | 2,627 (39%) | 353 (41%) | 310 (37%) | 1,037 (61%) | 55 (7%) |
| AV sclerosis | 1,396 (21%) | 148 (18%) | 203 (24%) | 0 (0%) | 274 (35%) |
| Mild AS | 2,000 (29%) | 263 (31%) | 237 (28%) | 209 (12%) | 313 (41%) |
| Moderate AS | 416 (6%) | 50 (6%) | 50 (6%) | 251 (15%) | 75 (10%) |
| Severe AS | 304 (5%) | 29 (3%) | 41 (5%) | 199 (12%) | 55 (7%) |
| **Age, years** | 68 (47–79) | 68 (46–78) | 69 (47–79) | 57 (41–70) | 77 (49–83) |
| **Male** | 3,401 (50%) | 415 (49%) | 424 (50%) | 842 (50%) | 390 (51%) |
| **Female** | 3,342 (50%) | 428 (51%) | 417 (50%) | 854 (50%) | 382 (49%) |
| **Body mass index, kg/m<sup>2</sup>** | 24 (22–26) | 24 (22–26) | 24 (22–26) | 23 (21–25) | 25 (22–27) |
| **LVEF, %** | 64 (60–67) | 64 (60–68) | 64 (59–68) | 66 (62–71) | 63 (59–67) |
| **Hypertension** | 726 (27%) | 77 (23%) | 87 (27%) | 412 (64%) | 96 (22%) |
| **Diabetes** | 527 (19%) | 81 (24%) | 51 (16%) | 179 (28%) | 78 (18%) |

Values are given as numbers (percentage) or median (interquartile range)
Abbreviations: AV, aortic valve; AVA aortic valve area; AS, aortic stenosis; LVEF, left ventricular ejection fraction.

### 3.2 View classification

Our view classification algorithm accurately identified the required images for assessing AS across all datasets. The overall accuracy rates were 99.6% for ITDS, 99.5% for DHDS, and 99.4% for TDDS. Detailed metrics are in ***Supplemental Results 1***.

### 3.3 Performance of DL-based AS continuum assessment algorithm

The DLi-ASc was calculated with an average processing time of less than 2 sec (1.8 ± 0.05 sec). The distribution of the DLi-ASc, produced by the DL-based AS severity continuum assessment algorithm, exhibited a consistent trend of increasing scores with the severity of AS across all datasets (***Figure 1a***). Interestingly, at the AV sclerosis stage, the DLi-ASc already significantly increased compared to the normal stage, indicating the algorithm’s ability to detect early changes. The DLi-ASc demonstrated an increasing trend as conventional parameters assessing AS severity, such as AV Vmax, mPG, and AVA, worsened (***Supplemental Results 2***). Using the stage-specific cutoffs derived from the validation dataset, we evaluated the diagnostic performance for identifying any AS, significant AS, and severe AS, with results summarized in ***Table 3***.

**Figure 1.**
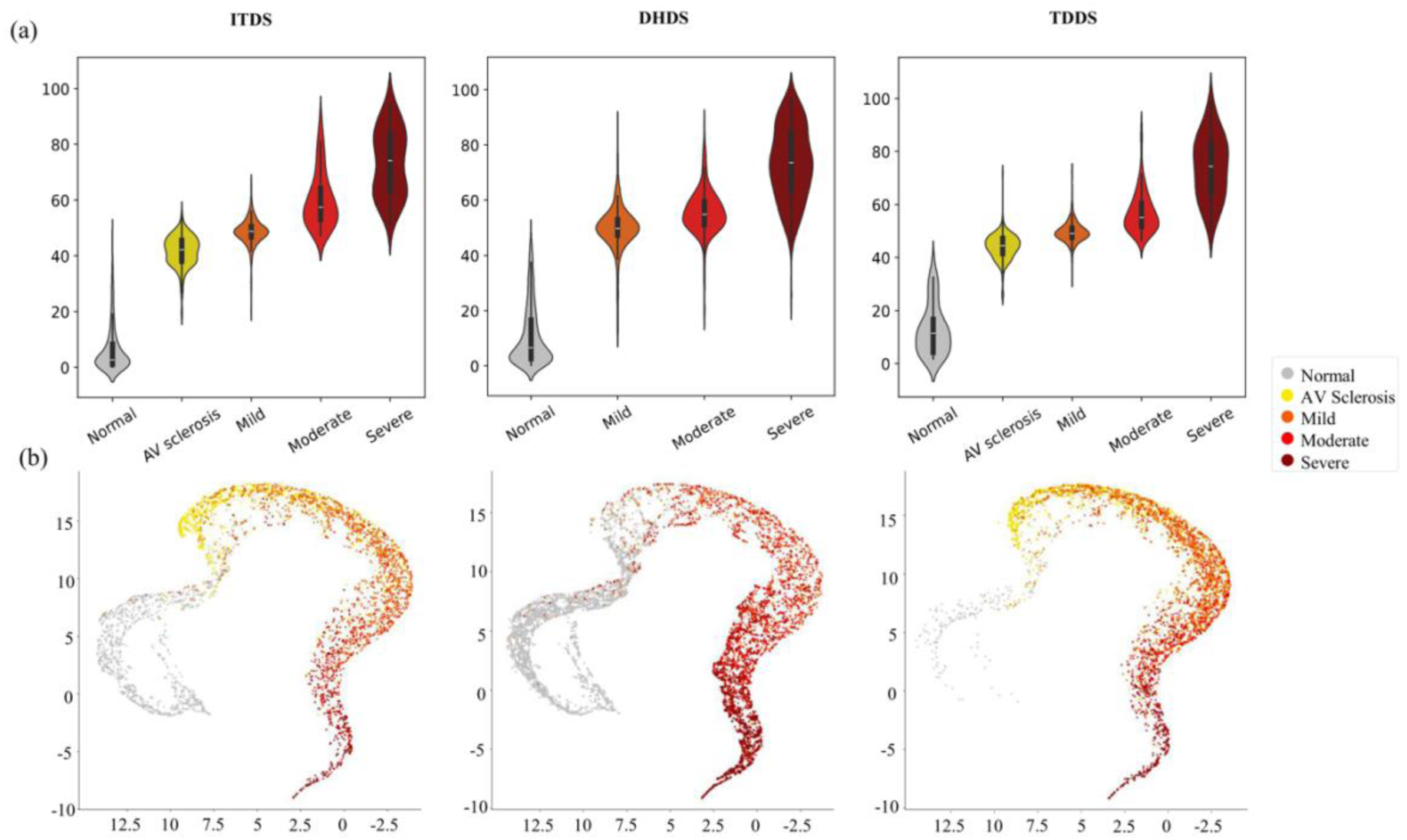
The Distribution of DLi-ASc According to AS Severity and UMAP Visualization. (a) The DLi-ASc, generated by the DL-based AS continuum algorithm, showed a consistent trend of increasing scores with the progression of AS severity observed across both internal and external datasets. (b) The UMAP plot demonstrates a continuous nonlinear gradient transition from the normal state (grey) through AV sclerosis (yellow) to advanced AS stages (red), visually underscoring the DLi-ASc accurately representing the AS continuum. Abbreviations as in Central Illustration: DHDS, distinct hospital dataset; ITDS, internal test dataset; TDDS, temporally distinct dataset; UMAP, uniform manifold approximation and projection.

**Table 3.**
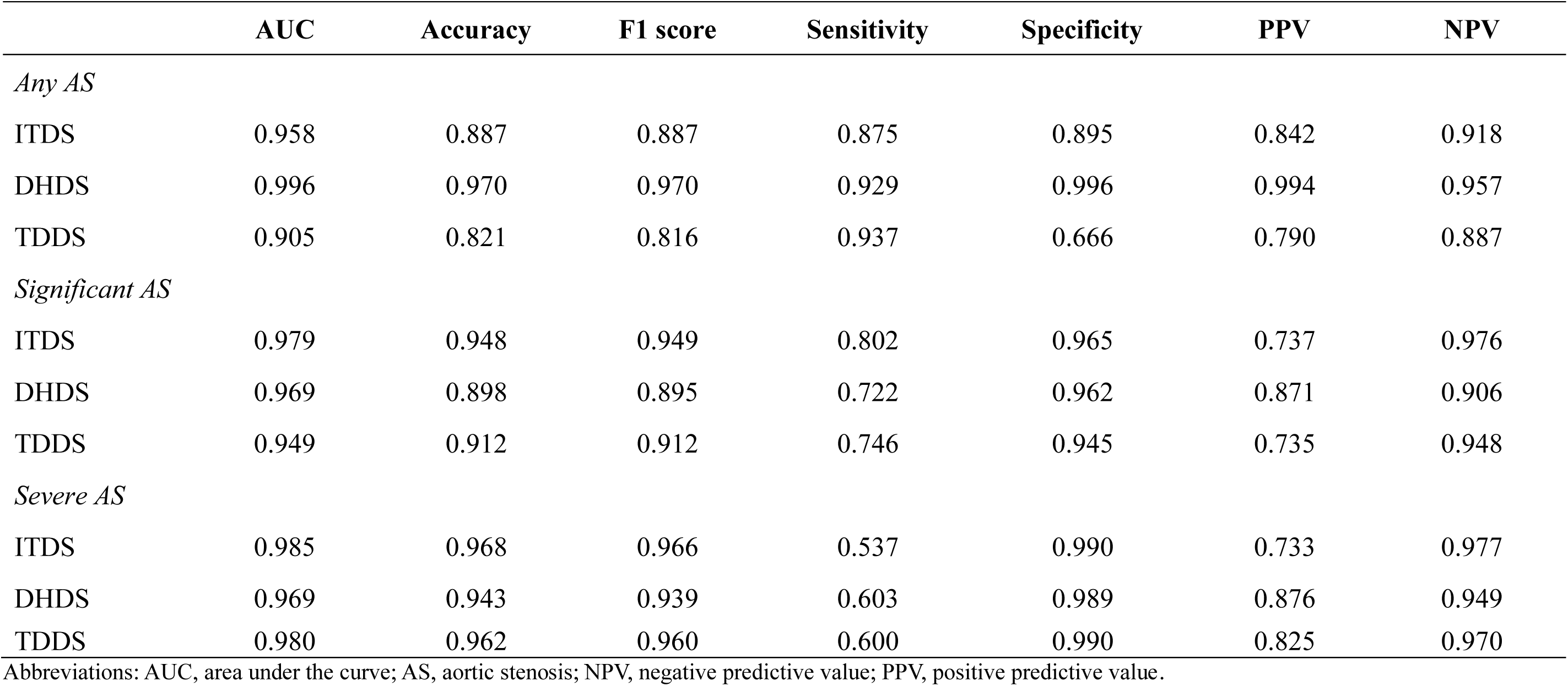
Diagnostic Performance of DLi-ASc Cutoffs for identifying Any AS, Significant AS, and Severe AS.

|  | AUC | Accuracy | F1 score | Sensitivity | Specificity | PPV | NPV |
| --- | --- | --- | --- | --- | --- | --- | --- |
| *Any AS* |  |  |  |  |  |  |  |
| ITDS | 0.958 | 0.887 | 0.887 | 0.875 | 0.895 | 0.842 | 0.918 |
| DHDS | 0.996 | 0.970 | 0.970 | 0.929 | 0.996 | 0.994 | 0.957 |
| TDDS | 0.905 | 0.821 | 0.816 | 0.937 | 0.666 | 0.790 | 0.887 |
| *Significant AS* |  |  |  |  |  |  |  |
| ITDS | 0.979 | 0.948 | 0.949 | 0.802 | 0.965 | 0.737 | 0.976 |
| DHDS | 0.969 | 0.898 | 0.895 | 0.722 | 0.962 | 0.871 | 0.906 |
| TDDS | 0.949 | 0.912 | 0.912 | 0.746 | 0.945 | 0.735 | 0.948 |
| *Severe AS* |  |  |  |  |  |  |  |
| ITDS | 0.985 | 0.968 | 0.966 | 0.537 | 0.990 | 0.733 | 0.977 |
| DHDS | 0.969 | 0.943 | 0.939 | 0.603 | 0.989 | 0.876 | 0.949 |
| TDDS | 0.980 | 0.962 | 0.960 | 0.600 | 0.990 | 0.825 | 0.970 |

Abbreviations: AUC, area under the curve; AS, aortic stenosis; NPV, negative predictive value; PPV, positive predictive value.

When discordant cases excluded from the training dataset were reintroduced in the ITDS, DLi-ASc for mild-to-moderate and LFLG moderate AS cases were positioned between mild and moderate AS, while moderate-to-severe and LFLG severe AS were distributed between moderate and severe AS (***Supplemental Results 3***). Evaluating DLi-ASc’s ability to differentiates LFLG severe AS cases from milder stages in DHDS and TDDS showed strong discriminatory performance. In DHDS, the AUC for distinguishing LFLG severe AS from normal to mild AS was 0.97, and from normal to moderate AS was 0.93. Similarly, in TDDS, the AUCs were 0.97 and 0.94, respectively, for these comparisons (***Supplemental Results 4***).

Furthermore, when we utilized UMAP to verify that the DLi-ASc accurately represents the AS continuum, the DLi-ASc, derived from the approach incorporating both ordered labels and multi-task learning, displayed a distinct continuous gradient from normal through AV sclerosis to advancing AS stages, consistently evident in ITDS and both external datasets (***Figure 1b***). In contrast, a conventional multi-class classification approach using 5-class cross-entropy loss resulted in the stage-based grouping but lacked the continuous progression seen in our approach. The continuous mapping with ordered labels approach, but without additional multi-task learning to predict key TTE parameters, appeared somewhat linear but did not accurately reflect the severity progression (***Supplemental Results 5***).

For each severity level, we present representative samples with Grad-CAM saliency maps overlaid on both PLAX and PSAX views, specifically localizing the AV (***Figure 2*** and ***Video S3***). These results demonstrate that our model accurately identifies the relevant regions for evaluating AS across all severity levels and views without supervision.

**Figure 2.**
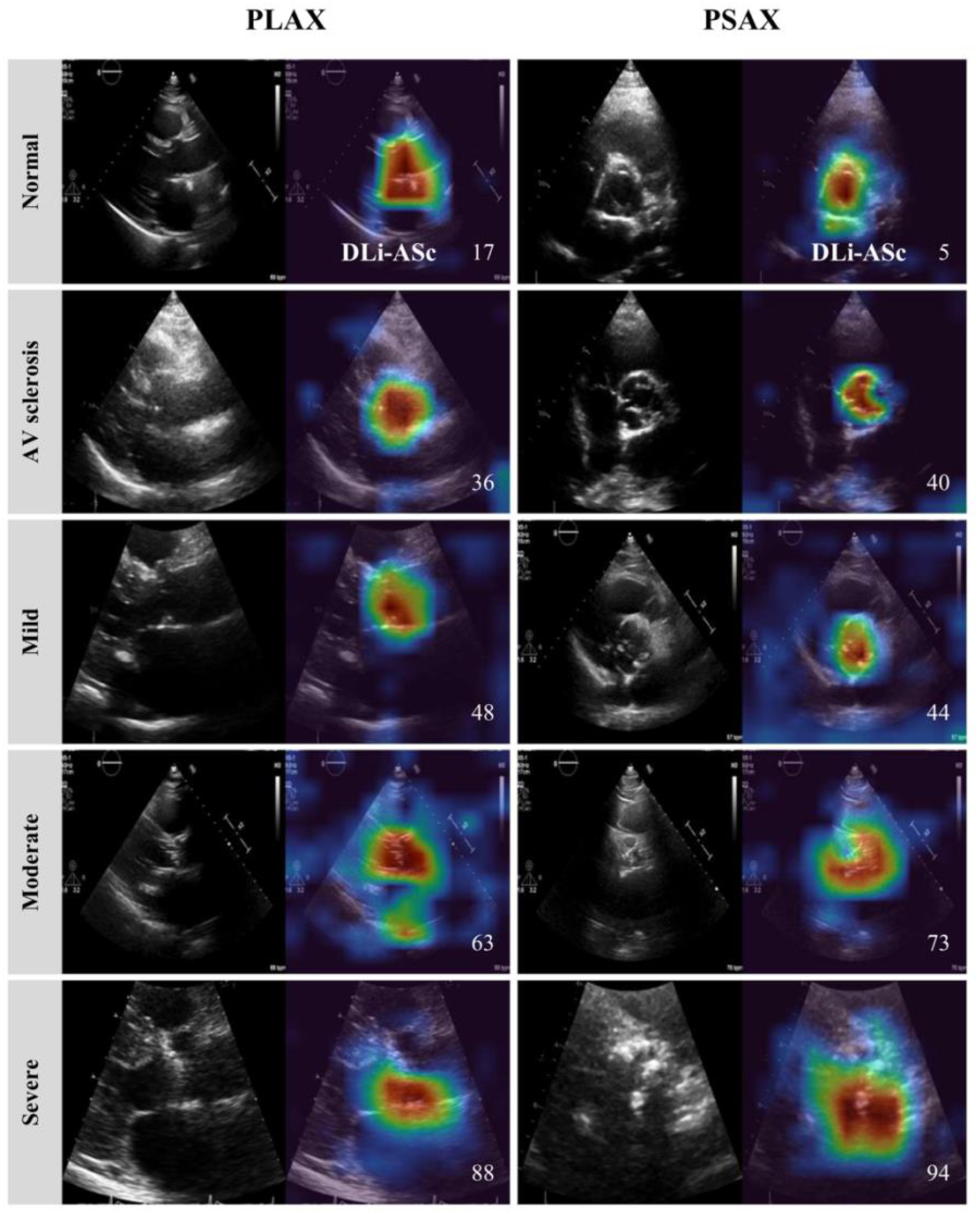
Explainability Analysis using Saliency Map. The figure displays representative PLAX and PSAX views alongside their corresponding Grad-CAM saliency maps during DLi-ASc calculation. The saliency maps highlight that the DL model accurately focuses on the AV. As AS severity progresses from normal to severe AS, the model produces increasingly higher DLi-ASc scores, corresponding worsening AS. Abbreviations as in Central Illustration: AV, aortic valve; PLAX parasternal long-axis view; PSAX, parasternal short-axis view.

### 3.4 Performance of automated conventional assessment algorithm

For conventional AS evaluation, measurements of spectral Doppler, including AV CW Doppler and LVOT PW Doppler, as well as LVOT diameter, are required. On average, each of these measurements took less than 0.5 sec (0.2 ± 0.02 sec) and less than 2.0 sec (1.4 ± 0.13 sec), respectively. Among patients with AS and available GT values, while some cases did not undergo automatic measurement due to high uncertainty, in most instances, our algorithm successfully performed automatic measurements of AV Vmax (100% success rate) and mPG (99.3–100% success rate) (***Supplemental Methods 8***). These measurements demonstrated strong correlations with the GT values for AV V_max_ (r 0.962–0.974; MAE 0.08–0.14 m/s) and mPG (r 0.958–0.971; MAE 1.23–2.82 mmHg) (***Figure 3a***). Besides, the algorithm successfully measured the LVOT diameter from PLAX videos, demonstrating robust concordance with manual measurement (r 0.618–0.738; MAE 0.10–0.11 cm). The correlation for AVA, calculated from these measurements, was also good (r 0.807–0.859; MAE 0.18–0.18 cm^2^) but relatively lower than V_max_ and mPG, due to its dependence on multiple measurements. Missing GT values resulted in fewer comparison cases (***Supplemental Methods 8***), and accumulated differences affected the overall accuracy. Nonetheless, the accuracy of AS severity classification based on these automated measurements remained strong (98.2% for ITDS, 82.1% for DHDS, and 96.8% for TDDS) (***Figure 3b***). We reviewed cases where there were large discrepancies between the GT and automatic measurements, and we have provided representative cases in ***Supplemental Results 6*.**

**Figure 3.**
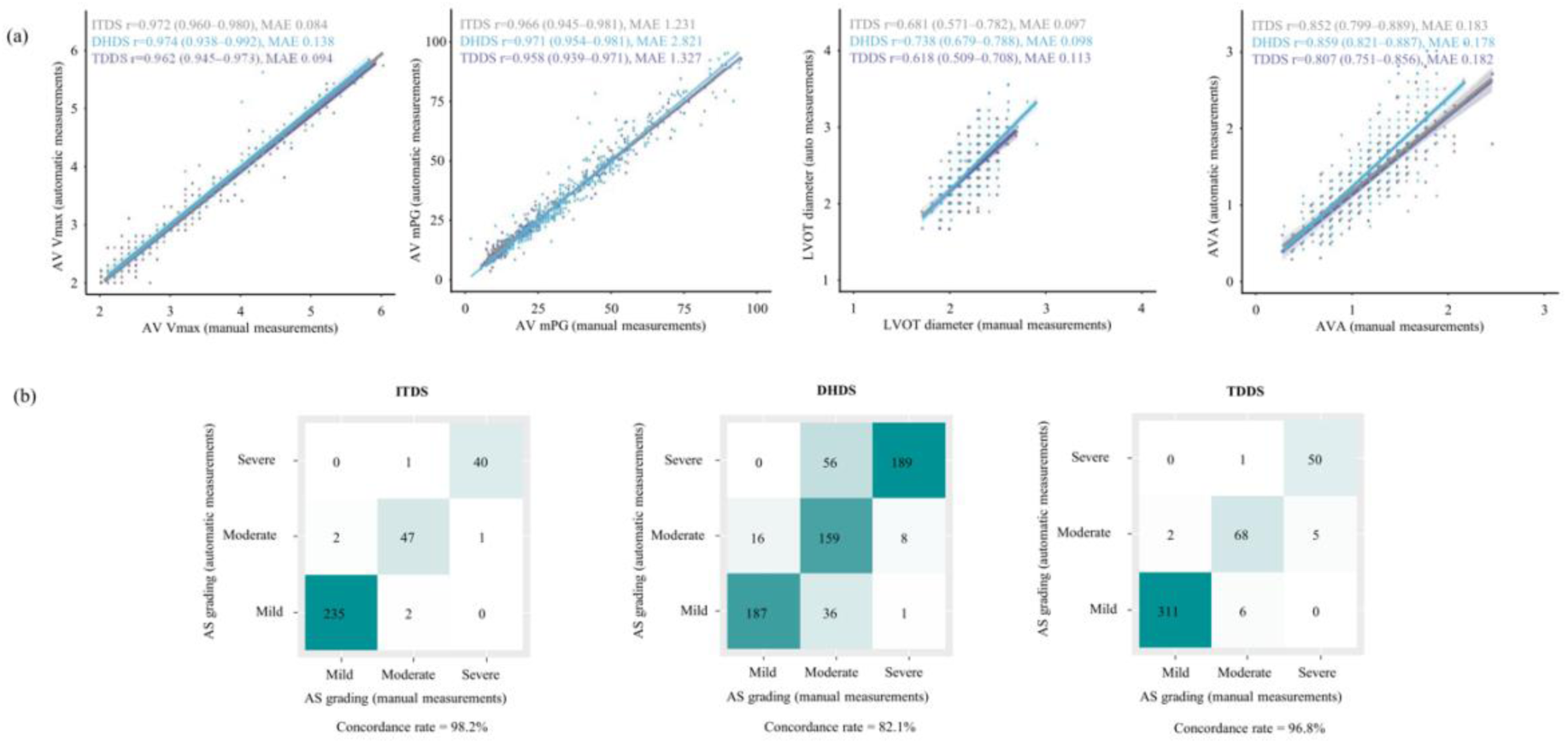
Concordance in AS Diagnosis Between DL-based Automated Assessment and Conventional Evaluation. (a) Across all datasets, the auto-measured AS parameters (AV maximal velocity, mean pressure gradient, and valve area) strongly correlated with those obtained from manual measurements. (b) Consequently, AS gradings from both methods exhibited a high concordance rate, ranging from 82.1% to 96.8%. Abbreviations as in Figure 1, 2 and 3: AVA, aortic valve area; LVOT, left ventricular outflow tract; MAE, mean absolute error; mPG, mean pressure gradient; r, Spearman Correlation Coefficient; V_max_, maximal velocity.

### 3.5 Comparison of diagnostic performance of two different AI-based approach

The discrimination performance of DLi-ASc for various stages of AS was generally excellent: AUC 0.91–0.99 for any AS, 0.95–0.98 for significant AS, and 0.97–0.99 for severe AS (***Figure 4***). When compared to automatically measured conventional parameters, in ITDS, the discrimination performance of DLi-ASc was lower than that of automatically measured V_max_ and mPG but comparable to AVA. In DHDS, the performance of DLi-ASc surpassed AVA in diagnosing all stages of AS, while in TDDS, it was comparable to AVA in diagnosing all stages of AS. Furthermore, when comparing DLi-ASc’s diagnostic performance with those of algorithms from previous studies on AS evaluation, DLi-ASc showed consistently superior accuracy in detecting any AS, significant AS, and severe AS across all datasets. The comprehensive results are available in **Supplemental Results 7.**

**Figure 4.**
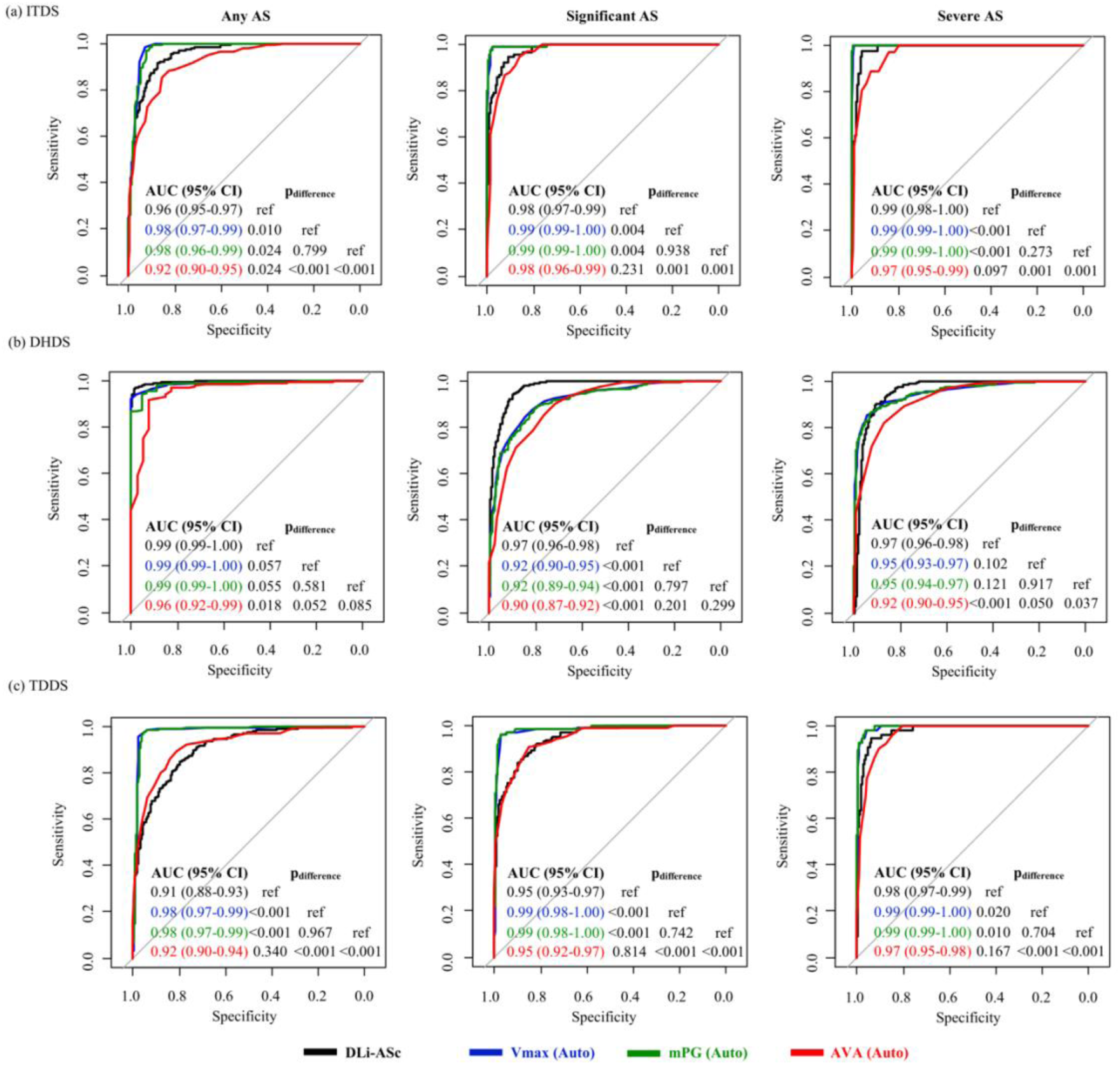
Diagnostic Performances of DLi-ASc and Other AI-derived Conventional AS Parameters Across Various Stages. The discriminative ability of DLi-ASc and other conventional AS parameters was consistently excellent for diagnosing any AS, significant AS (moderate to severe), and severe AS across all datasets: (a) ITDS, (b) DHDS, and (c) TDDS. Abbreviations as in Figures 1 and 3: AUC, the area under the curve; DLi-ASc, DL index for the AS continuum.

### 3.6 Prognostic value of AI-based AS assessment

Across median follow-up period [ITDS: 956 (278-1730) days, DHDS: 1435 (356-2140) days, and TDDS: 449 (174-563) days], 48 (15%), 349 (53%), 38 (9%) events were observed for ITDS, DHDS, TDDS, respectively. Analysis of spline curves across the ITDS, DHDS, and TDDS showed that an increase in DLi-ASc correlated with a rising risk of adverse clinical outcomes (***Figure 5***). The multivariable Cox regression analysis affirmed the strong and independent prognostic value of DLi-ASc. A 10-point increase in DLi-ASc from limited TTE videos was associated with hazard ratio (95% confidence interval) of 2.19 (1.77–2.71) in ITDS, and 1.64 (1.52–1.78) and 1.61 (1.31–1.99) in DHDS and TDDS, respectively (***Figure 6***). Moreover, the AI-derived parameters, such as V_max_, mPG, and AVA, demonstrated prognostic values comparable to those of manually-derived parameters (***Figure 6***).

**Figure 5.**
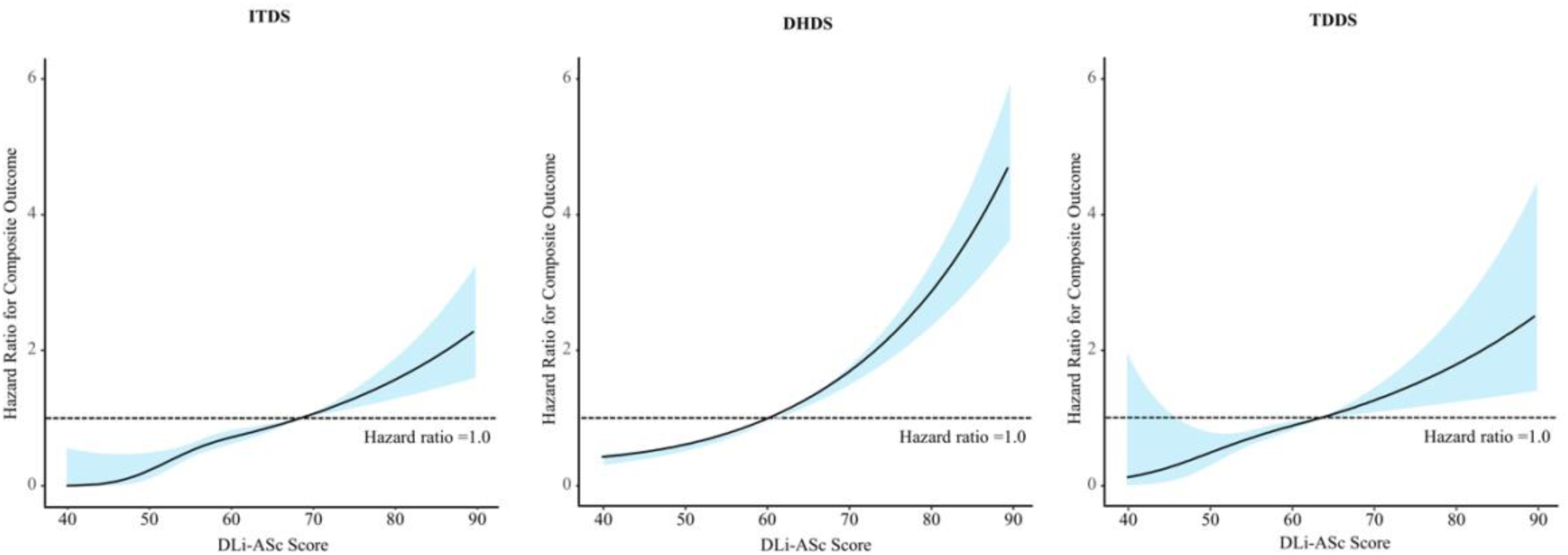
Spline Curves for Composite Outcomes Associated with DLi-ASc. The risk of composite outcome gradually increased with higher DLi-ASc across all datasets: (a) ITDS, (b) DHDS, and (c) TDDS. The solid lines represent the hazard ratio, and the blue shaded area represents the 95% confidence interval. Abbreviations as in Figures 1 and 3.

**Figure 6.**
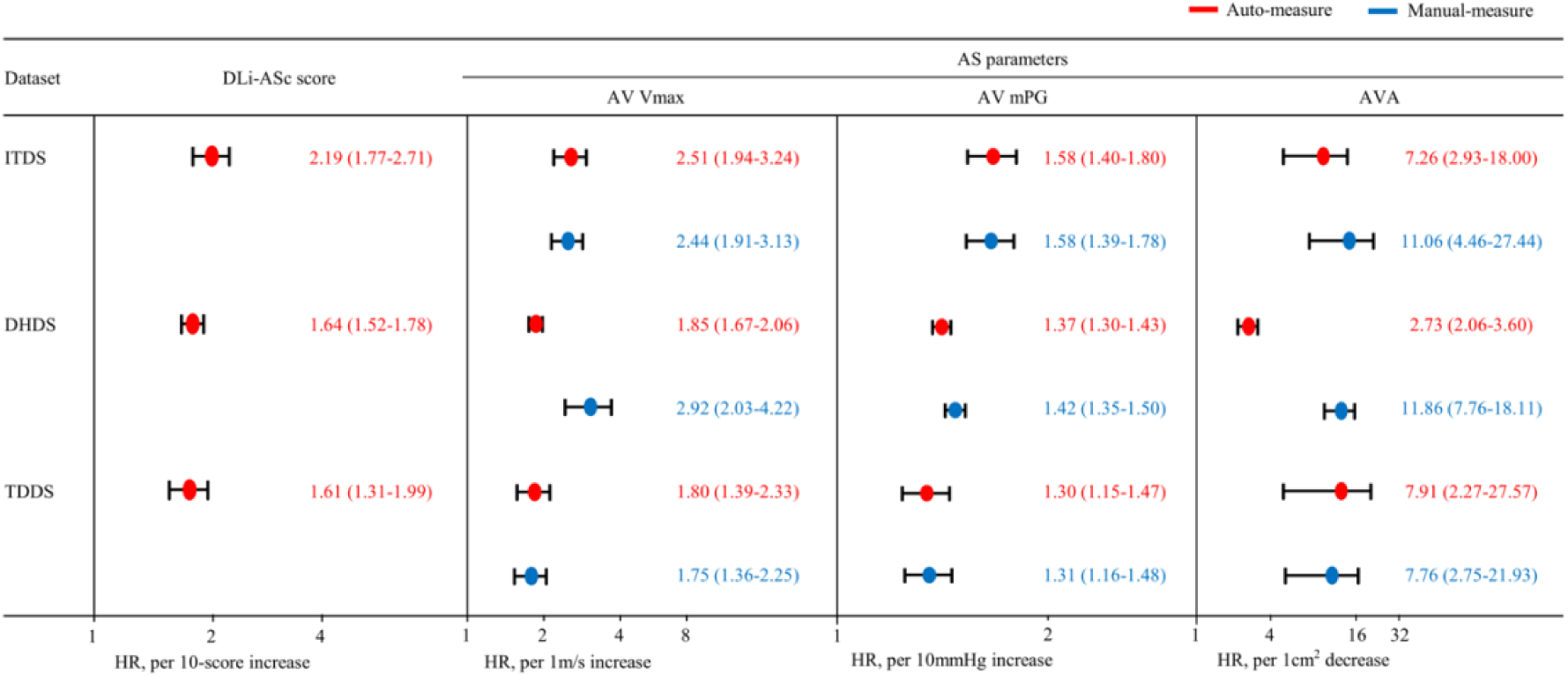
Prognostic value of DLi-ASc and AS parameters. The DLi-ASc showed independent predictive value for composite outcomes. Similarly, other AI-derived AS parameters were significant predictors for composite outcomes as well as manually-derived AS parameters. Abbreviations as in Figures 1, 2 and 3: HR, hazard ratio

## 4. Discussion

We have developed and validated a comprehensive AI-based system to evaluate AS through a dual pathway: 1) assessing the presence and severity of AS using only the PLAX and/or PSAX videos typically acquired early during TTE, and 2) automatically analysing additional views for conventional quantitative AS evaluation if obtained. This dual approach enables accurate AS evaluation across various settings, with internal and external validation demonstrating excellent diagnostic accuracy and strong prognostic capabilities.

While our AI-based system is not the first to evaluate AS, it stands apart from previous studies in several key aspects. First, our system provides both AS evaluation using limited 2D TTE videos and automation of conventional measurements. Prior research has typically focused on one of these aspects. For instance, Krishna et al. developed an AI model to automate quantitative AS evaluation.^5^ However, their model did not include the crucial initial visual analysis of the AV from 2D TTE videos, which is essential for initiating conventional quantitative AS analysis. Several studies used CNNs to extract AS-related features from 2D TTE videos through end-to-end learning without requiring Doppler information.^3,4,19,20^ Although these studies achieved decent performance in classifying AS severity, they lack conventional evaluation of AS, compromising trustworthiness, explainability, and interpretation. In contrast, our system integrates both approaches, identifying the potential for significant AS using parasternal views typically acquired early in TTE, guiding the acquisition of additional images for conventional AS evaluation, and providing automated analysis of these views. This approach not only predicts AS presence and severity from parasternal views, as human experts do, but also reduces workload by automating the subsequent conventional evaluation.

Another key strength of our study is that, unlike previous research, it reflects the continuous nature of AS progression. For instance, Wessler et al. trained CNNs to classify AS severity into three categories (no, early, and significant AS) using limited 2D images.^4^ Similarly, Ahmadi et al. proposed a transformer-based spatiotemporal architecture to classify AS into four categories (normal, mild, moderate, and severe AS) by capturing anatomical features and AV motion.^19^ Vaseli et al. focused on model explainability in AS severity classification, incorporating uncertainty estimation and classifying AS severity into three classes (no, early, and significant AS).^20^ However, these classifiers discretize AS severity, losing the continuum information of AS. Recently, Holste et al. proposed a binary classifier based on the 3D-ResNet18 architecture to detect severe AS, observing that model probabilities generated increase with AS severity.^3^ However, this model focused only on a binary classification task (e.g., non-severe vs. severe), not capturing the full range of AS severity levels in the training stage. In contrast, our framework employs continuous mapping with ordered labels, providing a more nuanced representation of AS severity. Importantly, we use multi-task learning with auxiliary tasks to predict continuous AS-related TTE parameters. This approach not only transitions from discrete labels to continuous values but also captures the underlying continuum of the disease more effectively. In UMAP visualizations, our model demonstrates a clear continuous gradient from normal to severe AS, unlike other classification models. Additionally, the appropriate distribution of DLi-ASc in discordant classification cases further supports for our framework: for instance, the DLi-ASc for mild-to-moderate AS cases was distributed between mild and moderate AS categories, while moderate-to-severe AS cases displayed DLi-ASc values between moderate and severe AS categories. This graded distribution aligns with the expected AS severity continuum, underscoring the algorithm’s ability to accurately assess AS severity even in discordant cases. Our comparative analysis with prior AI algorithms for AS evaluation^3,4^, which primarily employed binary or limited-category classifications, further supports the superiority or our approach, underscoring its robustness and potential clinical applicability.

Another value of this study lies in demonstrating the independent prognostic potential of the DL-derived novel index. While Strange G. et al. identified AS phenotypes linked to mortality, their model relied on tabular data from echocardiographic reports rather than direct image analysis.^21^ Conversely, Oikonomou EK et al. developed a video-based AI biomarker, the Digital AS Severity index (DASSi), similar to our study.^22^ DASSi (range 0–1) primarily predicted AV Vmax change per year and AVR risk, showing a 4–5-fold increased AVR risk for scores ≥0.2. Our study, however, derived the DLi-ASc (range 0–100), reflecting the AV severity continuum, where a 10-point increase in DLi-ASc corresponded to a 1.6–2.2-fold rise in the composite endpoint risk, including mortality, HF admission, and AVR. These findings reinforce the potential of image-based digital markers for prognostication in AS management.

The implications of our AI-based system extend beyond the reliable detection of significant AS; it also can provide guidance for less experienced operators and opportunities to identify and correct potential errors. This functionality mirrors the role of human expert’s visual evaluation in real-world practice. For example, the system can accurately predict the presence and severity of significant AS using only PLAX or PSAX videos, guiding operators to acquire additional necessary images, which can then be used for automatic quantitative AS evaluation. If image acquisition is suboptimal, as in the case of improperly acquired AV CW Doppler, the AS severity may be underestimated. Additionally, even when AV CW Doppler is properly acquired, inexperienced operators may struggle with the accurate interpretation of LFLG severe AS. In such cases, a high DLi-ASc could prompt a re-evaluation, accounting for potential errors in image acquisition or analysis. This expectation shaped our design of DLi-ASc to function similarly to a human expert’s initial visual assessment; thus, it provides assessment for all images regardless of image quality. In contrast, the automatic measurement process halts when high uncertainty or poor image quality is detected to prevent inaccurate automatic measurement. Despite these operational differences, DLi-ASc demonstrated high discriminatory performance, even for discrimination of LFLG severe AS. Nevertheless, further well-designed prospective studies are necessary to confirm whether DLi-ASc can reliably aid in cases where measurements are challenging or less dependable in clinical practices.Moreover, we found that the DLi-ASc increases significantly from normal levels at AV sclerosis and mild AS stages before significant AS progression, which provides additional value of our AI-based system. DLi-ASc is poised to effectively monitor AS progression from preclinical stages as a score-based tool. We anticipate the clinical utility of our system becoming prominent, especially as new pharmacological treatments are investigated for AS prevention are explored.^23,24^ If such treatments become available, our algorithm’s sensitivity in detecting early AS stages will be highly advantageous. For sure, further studies are needed to confirm whether DLi-ASc consistently increases in tandem with AS progression, which will be essential to expanding its clinical application.

The present study has some limitations. Although we developed and thoroughly validated our AI-based system using data from multiple centres, including internal and external validation, all the data were retrospectively obtained from tertiary centres in South Korea. Thus, caution is needed in its interpretation and clinical application. For instance, the diagnostic performance of DLi-ASc observed in this study may vary in cohorts with different pre-test probabilities. While the cut-off values presented here may serve as reference points, additional studies across diverse populations are essential before clinical application. Furthermore, all echocardiographic data used in this study were obtained from tertiary institutions, where skilled operators acquired TTE. It remains to be seen if the DLi-ASc will perform well on TTE videos acquired in truly resource-limited and novice settings. However, this also suggests that if PLAX or PSAX videos are adequately acquired, the DLi-ASc could potentially evaluate AS as accurately as in advanced settings. Additionally, for the clinical outcome analysis, the observational study design led to some patients being lost to follow-up, which could introduce selection bias related to incomplete follow-up duration. Therefore, further studies are required to confirm its diagnostic and prognostic performance in various clinical environments and among different populations and AS subtypes. We are planning additional validation studies in primary clinics and a multi-national study to address these concerns. Additionally, although we designed the DLi-ASc to reflect the AS severity continuum, it needs to be verified whether the DLi-ASc increases progressively with the natural progression of AS. This issue will be addressed in future studies.

We developed and validated a comprehensive AI-based system for evaluating AS. This system operates through a dual pathway: it assesses the presence and severity of AS using limited TTE videos and simultaneously automates conventional quantitative AS evaluation. Internal and external validations demonstrated excellent diagnostic accuracy and strong prognostic capabilities. While further validation in diverse clinical settings is necessary, our system is expected to enhance AS detection and evaluation in resource-limited settings or by novices, while simultaneously reducing workload in advanced settings.

## Data Availability

The AI-based frameworks utilized in this study were developed and validated using the Open AI Dataset Project (AI-Hub) dataset, an initiative supported by the South Korean government's Ministry of Science and ICT.

## Contributors

All authors contributed equally to this study. All authors have read and approved the final version of the manuscript. J.P, J.K, J.J, and Y.E.Y verified the underlying data of the current study.

## Data Sharing Statement

The AI-Hub data may be accessible upon proper request and after approval of a proposal. Data from TDDS cannot be made publicly available due to ethical restrictions set by the IRB of the study institution; i.e., public availability would compromise patient confidentiality and participant privacy. Please contact the corresponding author to request the minimal anonymized dataset. Researchers with additional inquiries about the deep learning model developed in this study are also encouraged to reach out to the corresponding author.

## Acknowledgements

This work was supported by a grant from the Institute of Information & communications Technology Planning & Evaluation (IITP) funded by the Korea government (Ministry of Science and ICT) (No.2022000972, Development of a Flexible Mobile Healthcare Software Platform Using 5G MEC); and the Medical AI Clinic Program through the NIPA funded by the MSIT. (Grant No.: H0904-24-1002). The funders had no role in study design, data collection and analysis, decision to publish, or preparation of the manuscript.

---

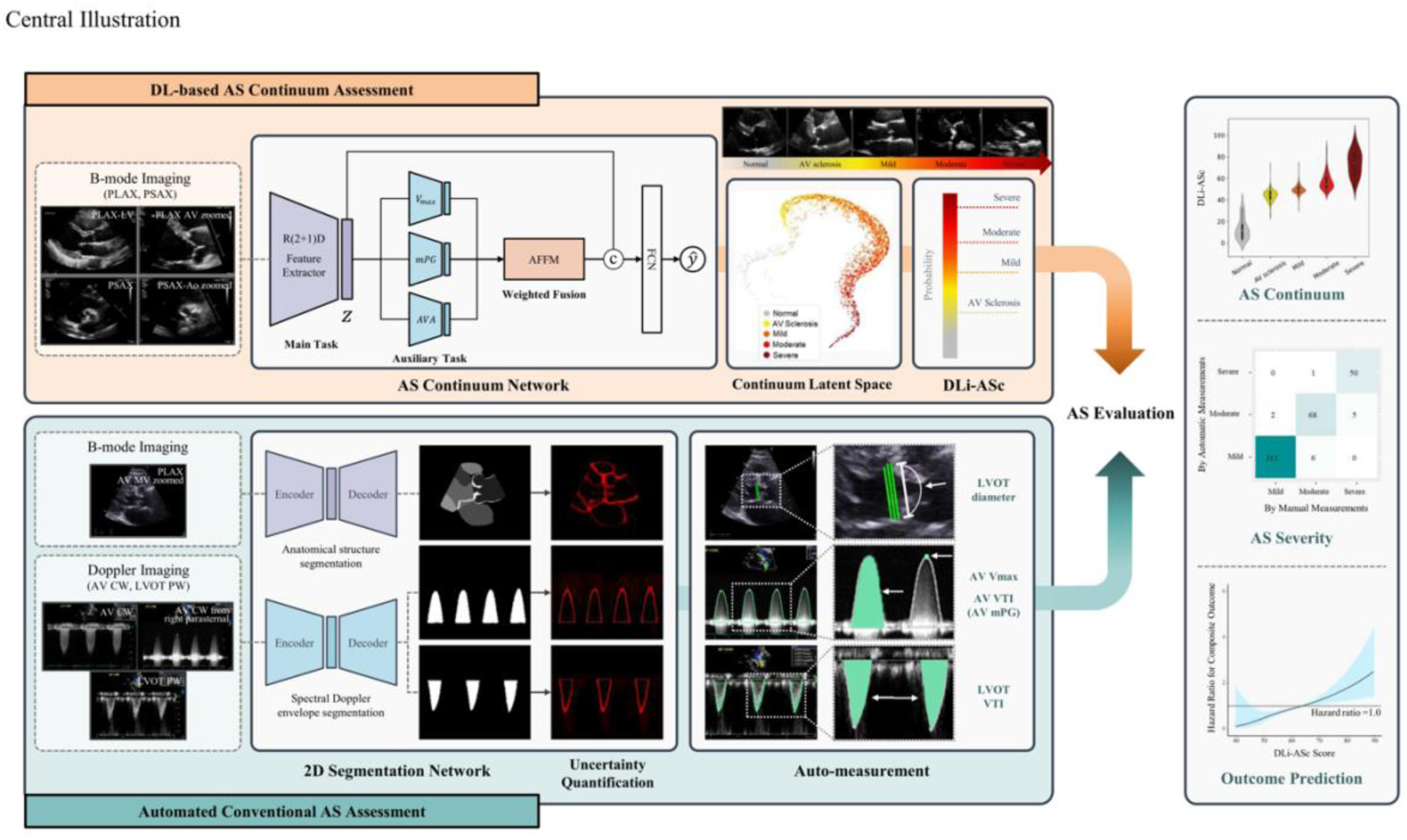
Central Illustration: AI-Enhanced Echocardiographic Assessment of AS Continuum. The illustration depicts a dual-pathway AI system for evaluating AS. The top row illustrates the DL-based assessment of the AS continuum using limited views, providing a unique DL index for the AS continuum, termed DLi-ASc. The bottom row demonstrates the automated AS assessment, which derives conventional echocardiographic AS parameters. By integrating both pathways, our AI system enables accurate AS diagnosis and prognostication, making it broadly applicable in advanced and resource-limited settings. AS, aortic stenosis; DL, deep-learning; DLi-ASc, DL index for the AS continuum.

