## Supplementary Material for "Artificial Intelligence-Enhanced Comprehensive Assessment of the Aortic Valve Stenosis Continuum in Echocardiography"

**Supplemental Methods 1. Study Population**

AI-Hub dataset consisted of 30,000 echocardiographic examinations retrospectively collected from five tertiary hospitals, including Chungnam National University Hospital, Hanyang University Hospital, Seoul National University Bundang Hospital, Severance Hospital, and Soonchunhyang University Seoul Hospital, over the period from 2012 to 2021. It encompasses a wide range of cardiovascular disease categories, from normal cases to ischemic heart disease, cardiomyopathy, pulmonary hypertension and embolism, pericardial disease, valvular heart disease, cardiac mass, and congenital heart disease. Data collection was based on diagnostic codes and echocardiographic reports to achieve targeted sample sizes; however, consecutive patient sampling was not enforced.

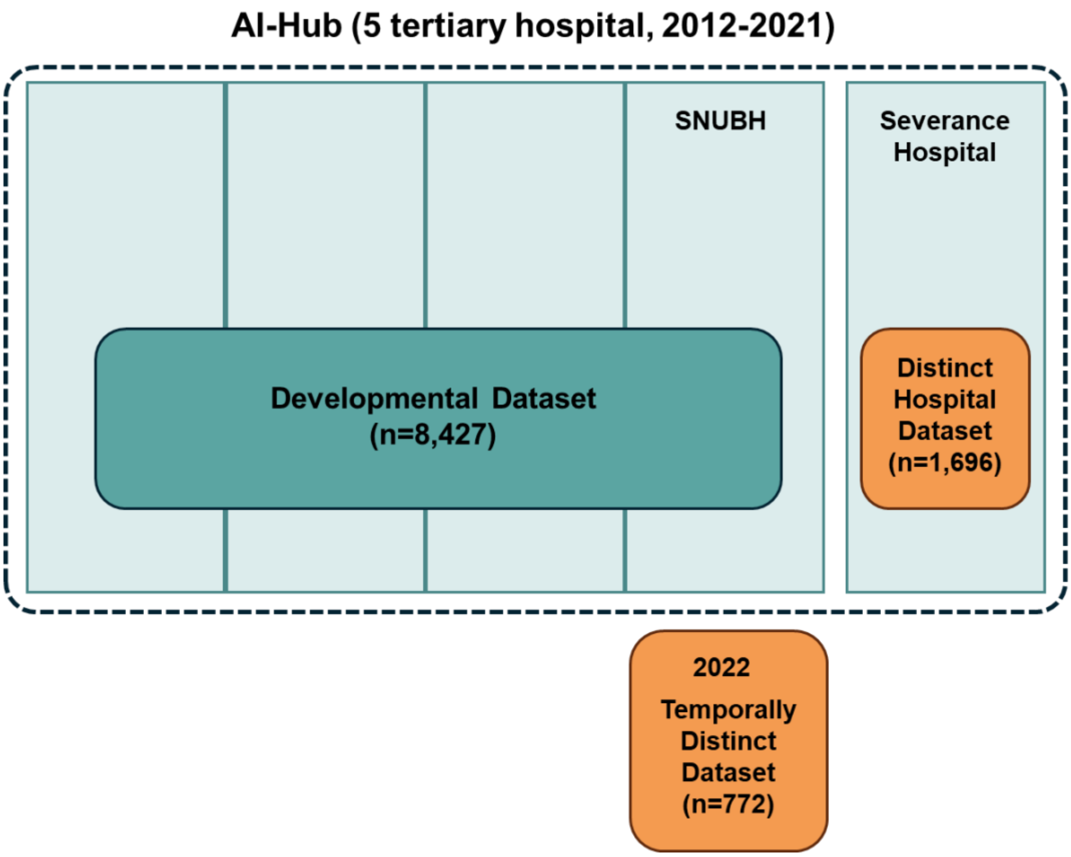


The AI-based frameworks introduced here were all developed using data extracted from the

AI-Hub dataset. Specifically, the DL-based AS continuum assessment algorithm was developed using the **Developmental Dataset (DDS)** sourced from the AI-Hub. During the assembly of DDS, data from Severance Hospital were deliberately excluded and used for external validation. We initially screened transthoracic echocardiography (TTE) data from 4,563 patients diagnosed with aortic stenosis (AS). After excluding those who had undergone aortic valve (AV) replacement or open-heart surgery, those with moderate or greater AV regurgitation, or cases where the severity of AS could not be determined, 4,018 AS patients have remained. To ensure the model's accuracy, 628 cases exhibiting discordant findings among aortic valve (AV) peak velocity ($V_{max}$), mean pressure gradient (mPG), and aortic valve area (AVA) regarding the severity of AS were excluded. These cases were later used separately for validating the model. Among the 3,390 AS-diagnosed patients included in the model development, 2,500, 516, and 374 were categorized into mild, moderate, and severe AS, respectively. Additionally, for the purpose of model training, TTE examinations were extracted for 3,290 individuals demonstrating normal AV morphology and function and 1,747 individuals exhibiting signs of AV sclerosis—characterized by degenerative changes in the AV but not meeting the diagnostic criteria for AS. Consequently, TTE data from a total of 8,427 individuals were compiled into the DDS. These data were split in an 8:1:1 ratio for training, validation, and internal testing purposes.

The **Distinct Hospital Dataset (DHDS)** was compiled by reviewing data from Severance Hospital that were not included in the DDS sourced from the AI-Hub dataset. A total of 719 AS patients were reviewed, none of whom had undergone AV replacement or open-heart surgery. After excluding 60 patients with moderate or greater AV regurgitation, the dataset included 659 AS patients (209 mild, 251 moderate, and 199 severe). Adding 1,037 normal patients, the DHDS totaled 1,696 patients. Since Severance Hospital does not commonly use the diagnosis of AV sclerosis, a separate AV sclerosis category was not included.

For the **Temporally Distinct Dataset (TDDS)**, we screened TTE data conducted in 2022 at Seoul National University Bundang Hospital, identifying 520 consecutive patients with AS. After excluding cases with a documented history of AV replacement or open-heart surgery, those with moderate or greater AV regurgitation, or cases where the severity of AS could not be determined, a total of 443 patients with AS remained (313 mild, 75 moderate, and 55 severe). Additionally, 55 individuals with normal AV and 274 with AV sclerosis, identified during the same period, were included, resulting in a total of 772 cases included for this dataset.

**Supplemental Methods 2. View Classification Networks Update**

To enhance our echocardiographic view classification algorithm, we expanded our datasets and refined the classification algorithm to include new views. Building upon our previous work, the dataset was broadened to encompass more granular classifications and additional views.[1] Specifically, we differentiated the parasternal long-axis (PLAX) zoomed view into four distinct categories: PLAX zoomed aortic valve (AV) (779 videos), PLAX zoomed mitral valve (MV) (279 videos), PLAX zoomed both AV and MV (1,357 videos), and PLAX zoomed aorta (502 videos). Additionally, we incorporated 663 CW Doppler AV images obtained from the right parasternal view. These enhancements are crucial for accurately measuring AV stenosis (AS) parameters, such as left ventricular outflow tract (LVOT) diameter, AV maximum velocity ($V_{max}$), mean pressure gradient (mPG), and AV area (AVA), which are critical for evaluating the severity of AS. The new data points were annotated using a Developmental Dataset (DDS), and the view classification network was subsequently retrained with this enriched dataset, employing the methodology previously detailed in our research.[1]

The tables clearly show the exact number of views used in training for each category, offering a comprehensive count for the latest version of our view classification algorithm.

**Target Views for Current Version of View Classification Algorithm.**

| Echo Mode | Echocardiographic View | Number of Views |
|---|---|---|
| B-mode | Parasternal long-axis left ventricle | 7,202 |
| | Parasternal long-axis zoomed AV | 779 |
| | Parasternal long-axis zoomed MV | 279 |
| | Parasternal long-axis zoomed AV & MV | 1,357 |
| | Parasternal long-axis zoomed aorta | 502 |
| | Parasternal short-axis, level of great vessels | 2,959 |

| | | |
|---|---|---|
| | Parasternal short-axis, level of mitral valve | 5,967 |
| | Parasternal short-axis, level of papillary muscle | 6,635 |
| | Parasternal short axis, level of apex | 7,143 |
| | Apical four-chamber | 4,647 |
| | Apical four-chamber zoomed left ventricle | 5,977 |
| | Apical four-chamber right ventricular-focused | 2,767 |
| | Apical five-chamber | 1,325 |
| | Apical two-chamber | 3,035 |
| | Apical two-chamber zoomed left ventricle | 5,708 |
| | Apical three-chamber | 2,535 |
| | Apical three-chamber zoomed left ventricle | 5,500 |
| | Subcostal four-chamber | 3,233 |
| | Subcostal long axis IVC | 2,717 |
| M-mode | M-mode through left ventricle | 3,879 |
| | M-mode through aorta and left atrium | 2,788 |
| | M-mode tricuspid annular plane systolic excursion | 708 |
| Spectral and tissue Doppler | PW Doppler mitral valve | 6,816 |
| | TDI mitral valve lateral annulus | 6,388 |
| | TDI mitral valve septal annulus | 705 |
| | CW Doppler mitral stenosis | 1,811 |
| | CW Doppler mitral regurgitation | 1,473 |
| | PW Doppler left ventricular outflow tract | 4,262 |
| | CW Doppler aortic valve | 3,864 |
| | CW Doppler aortic valve in parasternal | 663 |
| | CW Doppler aortic regurgitation | 1,094 |
| | CW Doppler tricuspid regurgitation | 8,910 |
| | PW Doppler right ventricular outflow tract | 2,935 |
| | CW Doppler pulmonic valve | 643 |
| | CW Doppler pulmonic regurgitation | 748 |
| | Pulmonary vein flow | 1,701 |
| | Total | 119,655 |

AV, aortic valve; CW, continuous wave; IVC, inferior vena cava; MV, mitral valve; PW, pulse wave; TDI, tissue Doppler imaging

**Supplemental Methods 3. DL-based AS Continuum Assessment Algorithm**

Given an input video, we used the r2plus1d architecture as a backbone.[2] Importantly, we modified the backbone network to avoid temporal down-sampling by maintaining a stride of 1 along the temporal axis. In addition to r2plus1d, we evaluated alternative backbone architectures, specifically r3d and mc3, to ensure that our model's performance was not dependent on a single feature extractor. The performance comparison among these backbones, detailed in the following Table, demonstrates consistent accuracy across configurations, with r2plus1d achieving competitive results.

**Comparisons analysis of backbone architecture**

| | mc318 | r3d18 | r2p1d18 |
|---|---|---|---|
| ITDS | | | |
| any AS | 0.956 | 0.959 | 0.952 |
| significant AS | 0.955 | 0.968 | 0.951 |
| severe AS | 0.971 | 0.974 | 0.972 |
| DHDS | | | |
| any AS | 0.996 | 0.997 | 0.997 |
| significant AS | 0.847 | 0.805 | 0.832 |
| severe AS | 0.897 | 0.890 | 0.899 |
| TDDS | | | |
| any AS | 0.902 | 0.898 | 0.899 |
| significant AS | 0.947 | 0.942 | 0.946 |
| severe AS | 0.977 | 0.979 | 0.982 |

From this feature $\boldsymbol{z}$, we employ four decoders, three of which are designed to predict continuous variables such as AV $V_{max}$, mPG, and AVA. Each auxiliary decoder is constructed to predict these continuous variables through regression. Each auxiliary decoder consists of two hidden layers with 512 units each, batch normalization and ReLU activation, followed by an output layer with a single unit and a Softplus activation function to ensure positive output values.

We then implemented a fusion module, termed the Adaptive Feature Fusion Module (AFFM), that generates a score for each continuous feature and then fuses them into one feature by a weighted sum of all features according to their scores. For features $z_{v_{max}}, z_{mPG}, z_{AVA}$ from the three auxiliary decoders before the output layer with a single unit, the fusion module computes a weight $w_{v_{max}}, w_{mPG}, w_{AVA}$ for each feature $z_{v_{max}}, z_{mPG}, z_{AVA}$ using a fully connected layer followed by batch normalization, ReLU activation, and a final fully connected layer with sigmoid activation. The fused feature $z^{fused}$ is then computed as a weighted sum of the features: $z_{fused} = w_{v_{max}} z_{v_{max}} + w_{mPG} z_{mPG} + w_{AVA} z_{AVA}$

The fused feature $z^{fused}$ is then concatenated with the original feature $z$ from the r2plus1d backbone, resulting in the final feature $z_{final}$: $[z; z_{fused}]$. The final classifier classifier processes this concatenated feature $z^{final}$ by linear function followed by a sigmoid function: $\widehat{y_{final}} = sigmoid(W z_{final} + b)$.

Input videos were resized to 224 x 224, and normalization was applied to a [0, 1] range for both training and inference. During training, if the number of frames exceeded the specified clip length (16 frames), a random start index was selected, motivated by the approach in Holste et al. If the number of frames was fewer, indices were evenly spaced to fit the clip length. For inference, the videos were divided into four intervals and stacked. If the total frames in the video are sufficient, the video is divided into multiple intervals with evenly spaced start indices. Each interval begins at a calculated start index, ensuring consistent spacing across the video's duration. A sequence of frames matching the clip length is then sampled from each start index, providing a balanced temporal coverage. If the frames were insufficient for multiple intervals but exceeded the clip length, the video was sampled, and the clip was replicated to match the intervals. If the frames were fewer than the clip length, indices were evenly spaced, and the clip was duplicated to match the intervals. The stacked inputs from the inference phase are averaged to enhance the model's generalization capabilities. The final DL index for AS continuum (DLi-ASc) was rescaled to 0-100 by multiplying the model output after the sigmoid function by 100.

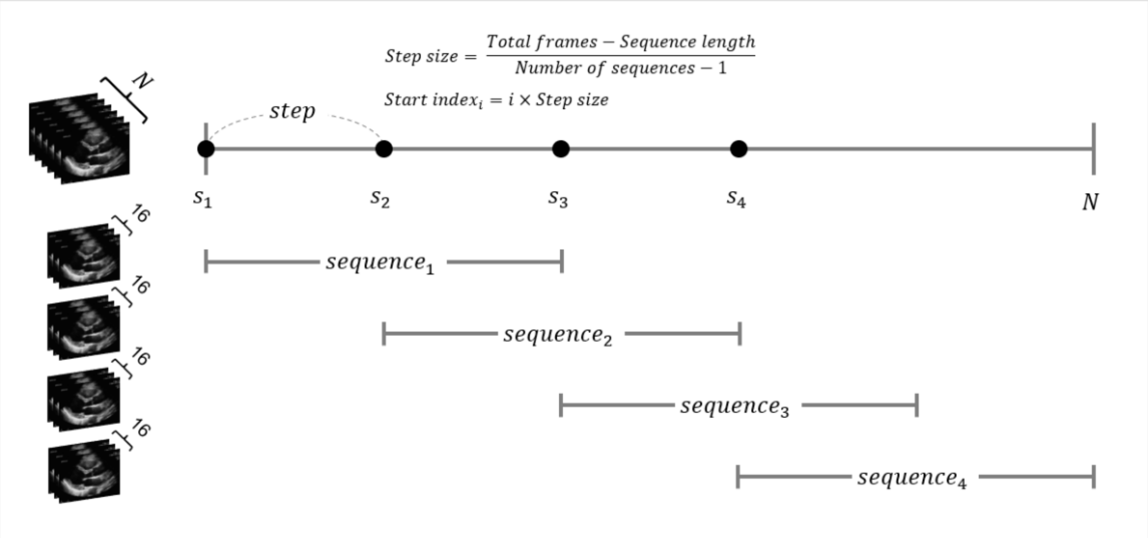


During training, a class sampler was used to balance the sampling of normal and AS classes by assigning higher weights to the less frequent class. The Adam optimizer was used with a learning rate of 0.0001 and a batch size of 28. No learning rate scheduler was utilized. Early stopping was implemented with a tolerance of 300 epochs, monitoring the validation loss as the metric. Additionally, in cases where ground-truth data for clinical variables such as AV $V_{max}$, mPG, and AVA were missing, the loss calculation for those instances resulted in a NaN value, which was excluded from the loss computation, ensuring that these instances did not affect model weight updates.

### Deep Learning Architecture for DLi-ASc

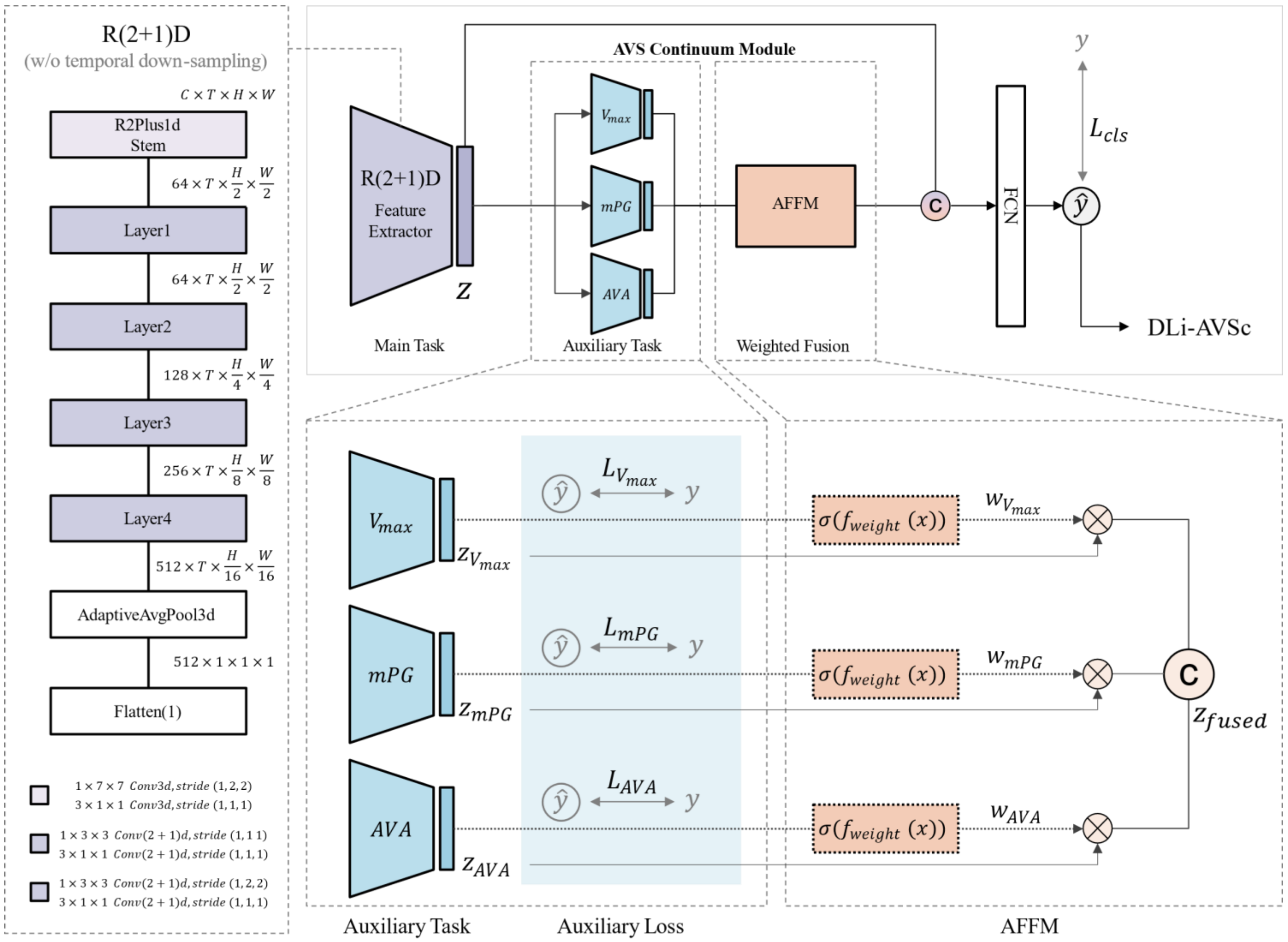

**Supplemental Methods 4. Cutoff of DLi-ASc for Each AS Severity Category**

We derived the DLi-ASc cutoffs by calculating the mean values of DLi-ASc across normal, AV sclerosis, mild, moderate, and severe AS groups in the validation dataset. To establish cutoff points, we took the midpoint between the mean DLi-ASc values of each consecutive AS severity category. For example, the mean DLi-ASc values for moderate and severe AS were 59.1 and 80.3, respectively, yielding a cutoff of 69.7 for diagnosing severe AS. This approach resulted in the following DLi-ASc cutoffs: 24.6 for AV sclerosis, 45.4 for mild AS, 53.7 for moderate AS, and 69.7 for severe AS.

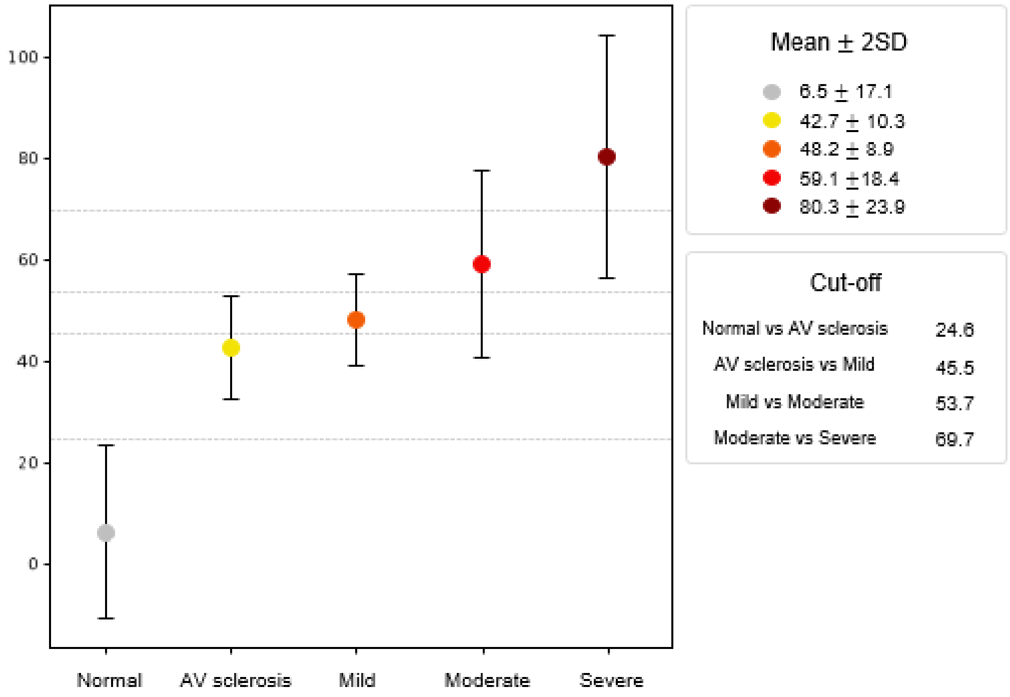

### Supplemental Methods 5. Automated Conventional AVA Assessment Algorithm

#### Supplemental Methods 5.1. Automatic Measurement of Spectral Doppler Echocardiography

The Doppler segmentation network employs the BiSeNetV2 architecture, which has been thoroughly described in our previous publication.[3-5] The BiSeNetV2 is specifically designed to balance accuracy and computational efficiency, making it suitable for real-time applications. During the training process, the network was optimized using standard cross-entropy loss. Notably, our Doppler segmentation network did not include training data for the AV continuous wave (CW) Doppler from the right parasternal view. However, this view can be inferred as a vertically flipped version of the CW Doppler AV in the apical view. During inference, we preprocess the input by flipping the image vertically before running it through the network, allowing us to utilize the same trained model for both views without additional training data.[5]

**AV CW Doppler from apical and right parasternal view**

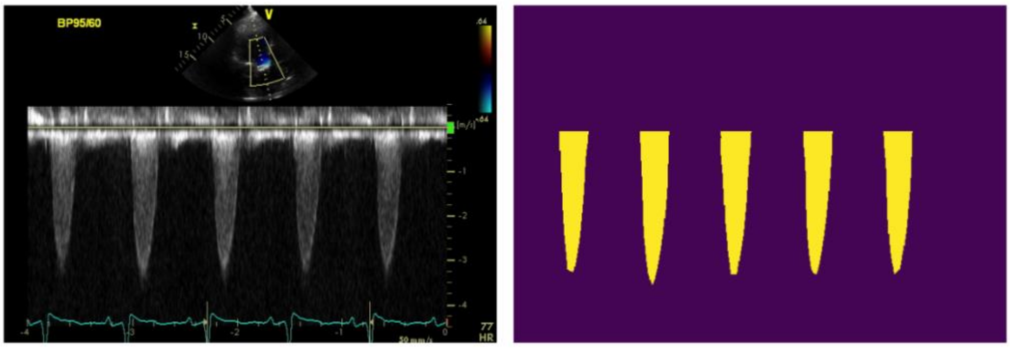


AV CW Doppler from Apical View

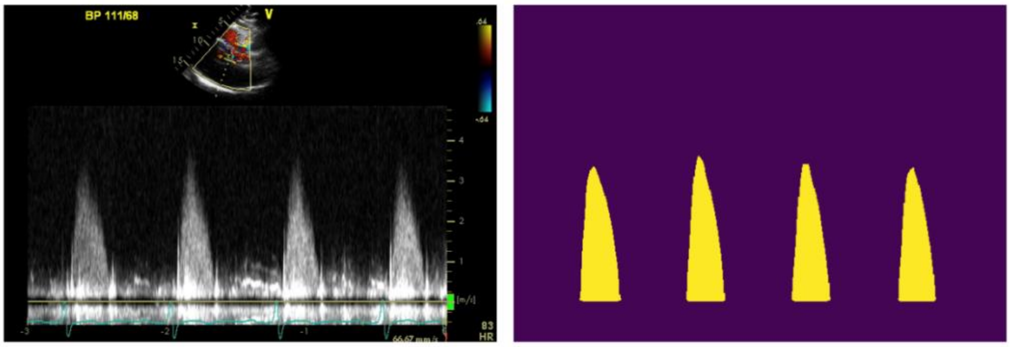


AV CW Doppler from Right Parasternal View

**Supplemental Methods 5.2. Segmentation Network for Parasternal Long-Axis (PLAX) View**

For the PLAX segmentation network, we utilized the SegFormer architecture, which includes a transformer encoder that provides multiscale features without needing positional encoding and a lightweight multi-layer perceptron (MLP) decoder integrating local and global attention for efficient segmentation.[6] A weighted cross-entropy loss was applied during training to account for the relatively small size of the mitral valve (MV) and AV in the PLAX view. A total of 2,369 PLAX videos were annotated at four key frames in the cardiac cycle: end-diastole, mid-systole, end-systole, and mid-diastole. Three experienced sonographers – MJ Jung and A Choi (each with 20 years in the field) and AR Kim (with 10 years) – performed the annotations. All segmentations were subsequently reviewed by SA Lee, a cardiologist specializing in echocardiography, with 10 years of experience. The images were resized into 512 × 512 and normalized to [-1,1]. We used the Adam optimizer with a learning rate of 0.001 and incorporated RandAug, enhanced with echocardiography-specific augmentations such as shadow, depth attenuation, and haze, to improve model robustness.[7,8] Additionally, a cosine annealing learning rate schedule was employed to optimize the training process.[9] Complete videos are presented in **Video S1**.

**Human Expert Annotation and AI Predicted Mask in PLAX View**

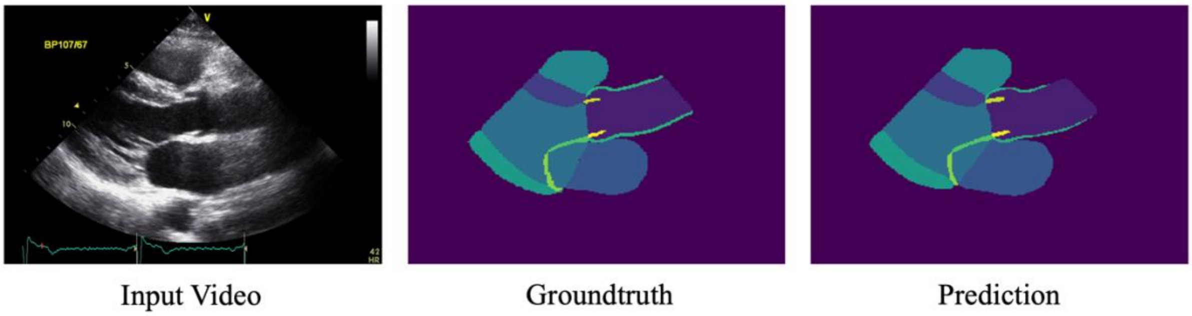

**Supplemental Methods 6. Quantifying Uncertainty in Predicted Segmentation**

Quantifying uncertainty in segmentation predictions is a meticulous process, crucial because segmentation errors can significantly impact the accuracy of subsequent automatic measurements. This uncertainty arises from two primary sources: epistemic uncertainty, which arises from a lack of knowledge of the DL model, and aleatoric uncertainty, which results from inherent noise in the data. To quantify these uncertainties, we calculate predictive entropy from the segmentation network's probability map, which provides a measure of the total uncertainty by combining both aleatoric and epistemic uncertainties. The entropy is computed for each pixel in the segmentation map, allowing us to identify regions with high uncertainty. The entropy is calculated using the equation:

$$H[\boldsymbol{p_{ij}}] = \sum_c p_{ij}^c \log p_{ij}^c,$$

where i, j represent pixel coordinates and c represents the class.

For quantifying uncertainty in the LVOT measurements in the PLAX view, we focus on regions of interests (ROIs) that directly affect the performance of LVOT measurement. Using the detected two points marking the annulus, we set a 50 × 50 ROI (10% of the resized image) centered on these points, as shown in the figure below. We then summed the entropy of each pixel within this ROI to assess uncertainty. For Doppler measurements, we evaluated uncertainty for the Doppler signal in each single beat. With the detected significant Doppler flow, we create an ROI and crop the entropy map to the corresponding ROI, normalizing it to 64 × 64. By summing the entropy values within the normalized ROI, we obtain the quantified uncertainty for each Doppler signal by beat. Complete videos are presented in **Video S2**.

### Regions of Interest marked for assessing uncertainty

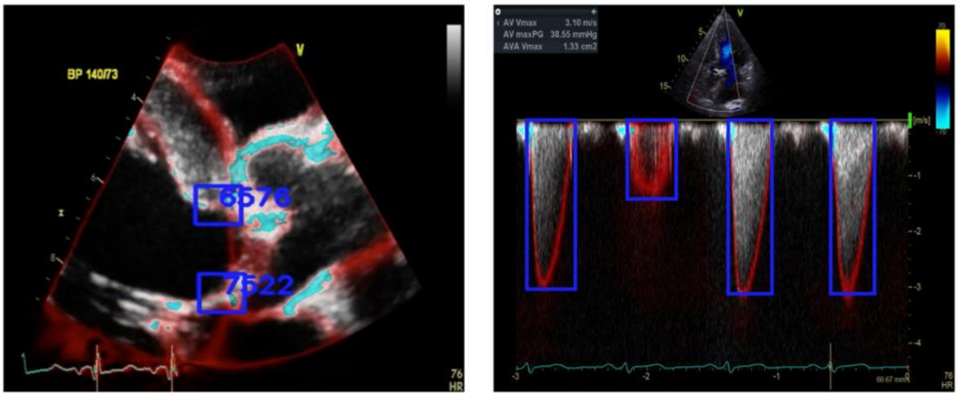


PLAX zoomed AV CW Doppler AV

From the validation set used for training, we find thresholds to reject the frames or beats by identifying the top percentage of frames or beats with the highest uncertainty. Specifically, we reject the top 5% of frames with the highest uncertainty from PLAX AV zoomed videos and the top 1% of beats with the highest uncertainty from Doppler images. We calculate the mean of entropy scores and add 1.96 × s.d. for PLAX frames and 2.33 × s.d. for Doppler images. The distributions of the entropy scores for both PLAX and Doppler images are shown in the figure below, illustrating how the thresholds are set.

### Distribution of Entropy Scores

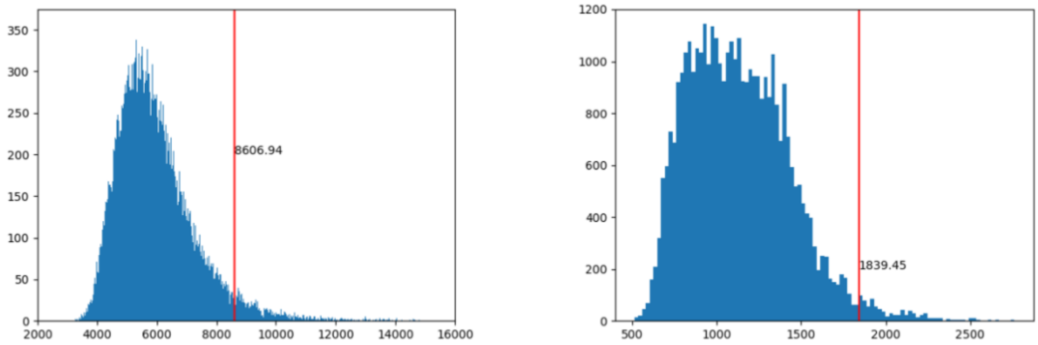


PLAX zoomed AV Doppler

**Samples with High Uncertainty Identified through Entropy Scores**

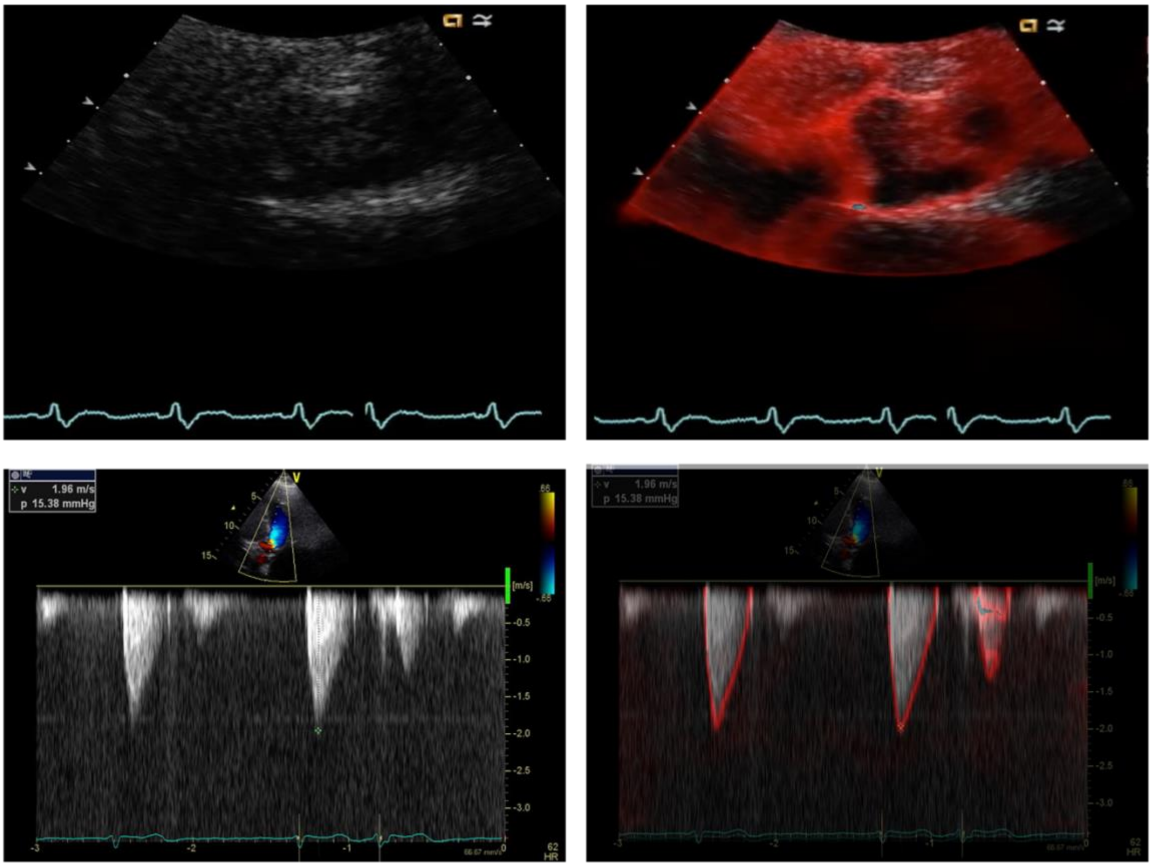


The figure above displays examples of Doppler and B-Mode images that were filtered out due to high uncertainty. These samples belong to the right end of the uncertainty distribution, representing the top percentage of frames and beats with the highest entropy scores. The highlighted regions in these images show areas of significant uncertainty, which were excluded from further analysis to ensure the accuracy of the automatic measurements. For spectral Doppler, the in/out filter was applied on a cardiac cycle basis. As seen in the AV CW Doppler image, the first two cycles were analyzed normally due to low uncertainty, while the last cycle was excluded due to high uncertainty.

**Supplemental Methods 7. Automatic Measurement of LVOT Diameter**

From the predicted segmentation mask, we identify points where the MV intersects with the aorta and where the septum intersects with the aorta to determine annulus points. Using these points, we measure the LVOT diameter at three locations: 1) at the annulus, 2) 2.5mm away from the annulus towards the LV cavity, and 3) 5mm away from the annulus towards the LV cavity. This approach reflects differing opinions on the appropriate location for measuring the LVOT diameter.[10] Complete videos are presented in **Video S1**.

**Automatic LVOT Measurements From the Predicted Segmentation Mask**

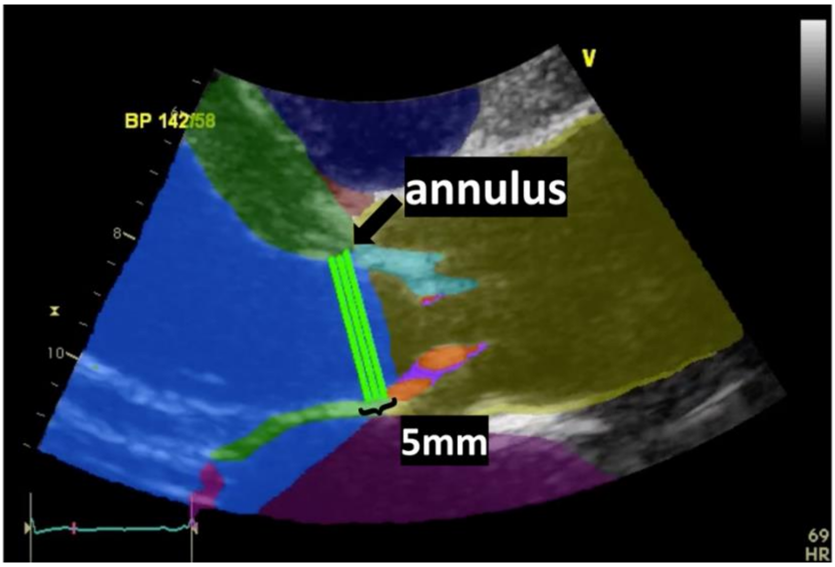

**Supplemental Methods 7.1. Validation of LVOT Diameter Measurement**

To assess the accuracy of the LVOT diameter automatic measurement, part of our AI-based system (Sonix Health, Ontact Health, Korea), we conducted a validation study with 212 American patients who underwent echocardiographic examination at Mayo Clinic Arizona, USA and Severance Hospital in Seoul, Korea. Considering the inherent variability in LVOT measurements, both AI and manual measurements were taken on the same view and frame. The AI system measured the LVOT diameter at three specific points: the annulus, 2.5mm from the annulus towards the LV side, and 5mm from the annulus, all marked in fluorescent green. Manual measurements were conducted at the annulus (dark red) and 5mm from the annulus (dark green) for comparison. The association between automated and manual measurements was assessed using the Spearman correlation analysis (r) and mean absolute error (MAE).

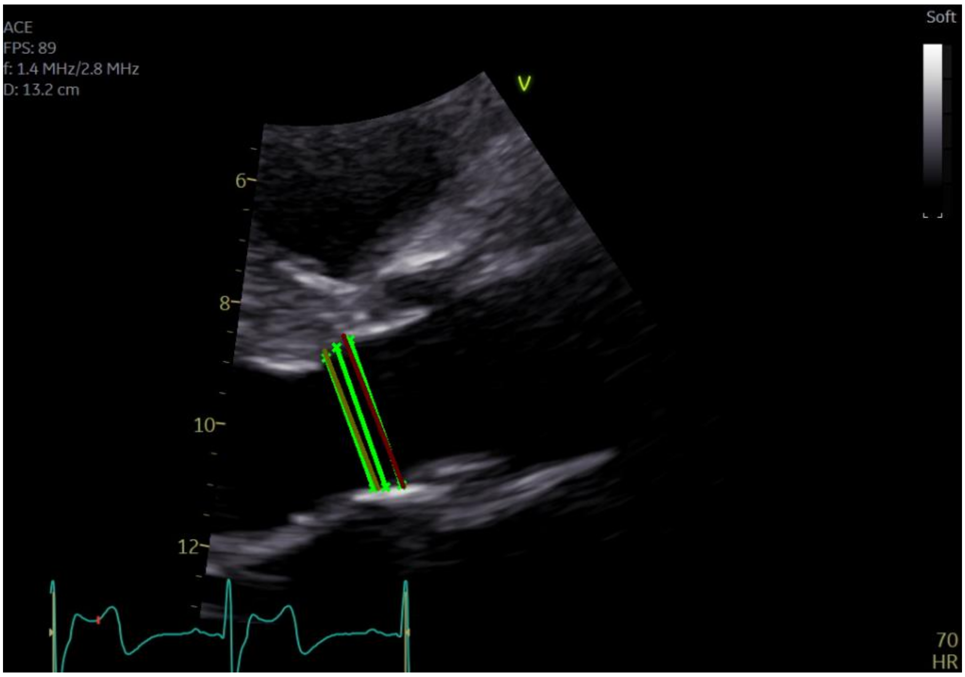

| **Comparison** | **LVOT Measurement Location** | **r** | **MAE (cm)** |
| --- | --- | --- | --- |
| Manual vs Manual | Annulus - 5mm from Annulus | 0.931 | 0.091 |
| Manual vs AI | Annulus - Annulus | 0.913 | 0.116 |
| | Annulus - 2.5mm from Annulus | 0.904 | 0.114 |
| | Annulus - 5mm from Annulus | 0.884 | 0.144 |
| | 5mm from Annulus - Annulus | 0.909 | 0.148 |
| | 5mm from Annulus - 2.5mm from Annulus | 0.948 | 0.114 |
| | 5mm from Annulus - 5mm from Annulus | 0.948 | 0.114 |

**Supplemental Methods 7.2. Selection of Optimal LVOT Diameter Measurement Location for AVA Calculation**

Our AI system provides LVOT diameter measurements at three locations. To determine which value to use for AVA calculation in this study, we compared the AI measurement at each location to prior manual measurements within the training set. Results showed that the automated measurement at the annulus had the highest correlation with manual measurement and lowest mean absolute error (MAE), leading us to select the annuus measurement for AVA calculation.

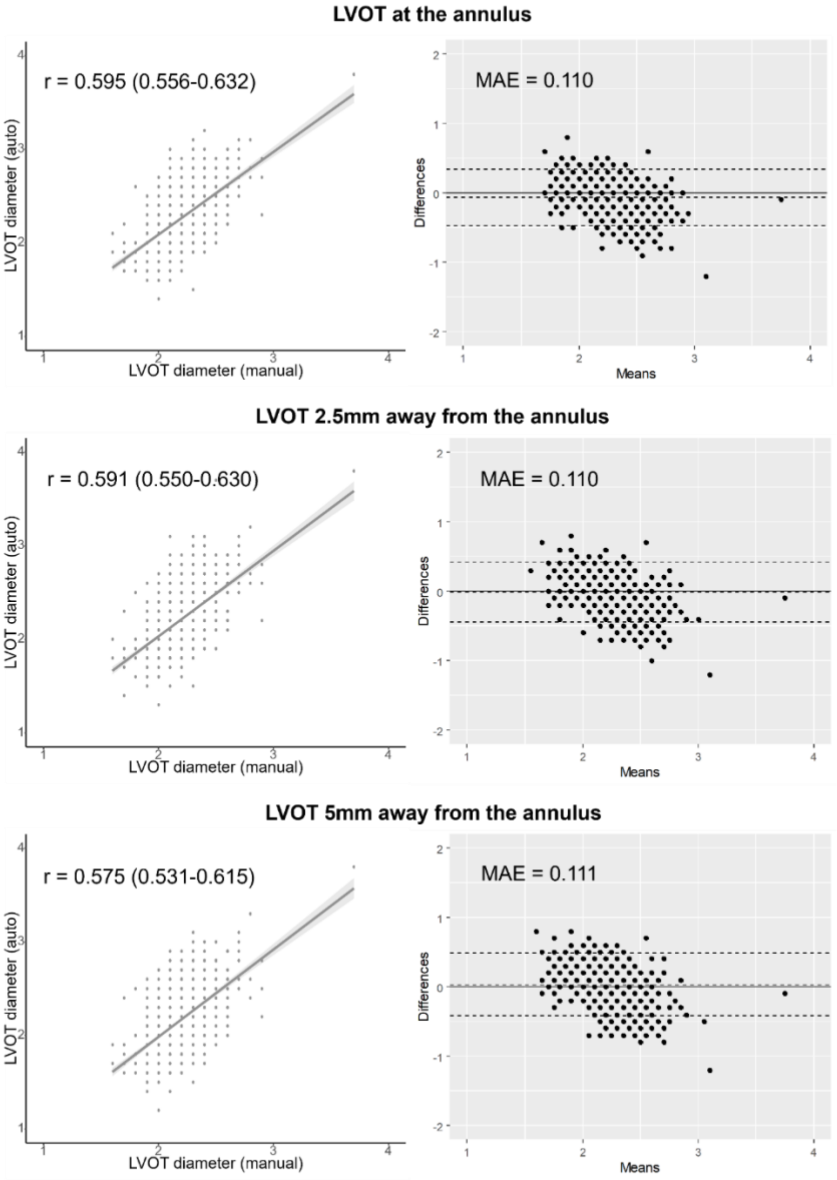

**Supplemental Methods 8. Availability of Ground Truth Measurements and Success Rate of Auto-Measurements in the AS Group**

The availability of ground truth measurements and the success rate of our algorithm's auto-measurements in the AS group are as follows.

| | **Ground truth (% of the overall case)** | **Auto-measurement (% of the overall case)** | **Matching case (% of available GT cases)** |
|---|---|---|---|
| *ITDS (n=328)* | | | |
| AV Vmax | 328 (100) | 328 (100) | 328 (100) |
| AV mPG | 320 (97.6) | 328 (100) | 320 (100) |
| LVOT VTI | 166 (50.6) | 164 (50.0) | 164 (98.8) |
| LVOT diameter | 159 (48.5) | 290 (88.4) | 141 (88.7) |
| AVA | 156 (47.6) | 143 (43.6) | 133 (85.3) |
| *DHDS (n=659)* | | | |
| AV Vmax | 83 (12.6) | 652 (98.9) | 83 (100) |
| AV mPG | 602 (91.4) | 652 (98.9) | 598 (99.3) |
| LVOT VTI | 438 (66.5) | 583 (88.5) | 367 (83.8) |
| LVOT diameter | 425 (64.5) | 618 (93.8) | 405 (95.3) |
| AVA | 560 (85.0) | 543 (82.4) | 457 (81.6) |
| *TDDS (n=443)* | | | |
| AV Vmax | 443 (100) | 443 (100) | 443 (100) |
| AV mPG | 440 (99.3) | 443 (100) | 440 (100) |
| LVOT VTI | 235 (53.0) | 233 (52.6) | 233 (99.1) |
| LVOT diameter | 228 (51.5) | 419 (94.6) | 212 (93.0) |
| AVA | 227 (51.2) | 219 (48.8) | 209 (96.5) |

AV, aortic valve; AVA, aortic valve area; LVOT, left ventricle outflow tract; mPG, mean pressure gradient; $V_{max}$, peak aortic valve velocity; VTI, velocity time integral.

**Supplemental Results 1. View Classification Performance in Each Dataset**

In this study, view classifications performance was assessed based on the framework's ability to accurately identify views necessary for AS evaluation. All other echocardiographic views are categorized as "Other", as they do not directly contribute to AS evaluation within our framework.

**ITDS**

| | n | Precision | Recall (Sensitivity) | specificity | F1-score | Accuracy |
|---|---|---|---|---|---|---|
| PLAX-LV | 1,525 | 0.985 (0.985, 0.989) | 0.991 (0.991, 0.995) | 1.000 (1.000, 1.000) | 0.988 (0.984, 0.992) | |
| PLAX zoomed AV | 197 | 0.985 (0.984, 1.000) | 0.685 (0.685, 0.711) | 0.999 (0.999, 0.999) | 0.807 (0.757, 0.852) | |
| PLAX zoomed AV & MV | 663 | 0.896 (0.896, 0.904) | 0.979 (0.979, 0.987) | 1.000 (1.000, 1.000) | 0.936 (0.924, 0.948) | |
| PSAX, level of great vessels | 1,819 | 0.990 (0.990, 0.994) | 0.971 (0.971, 0.974) | 0.999 (0.999, 0.999) | 0.980 (0.976, 0.985) | |
| PW Doppler LVOT | 886 | 0.965 (0.965, 0.971) | 0.998 (0.998, 1.000) | 1.000 (1.000, 1.000) | 0.981 (0.975, 0.987) | 0.996 (0.996, 0.996) |
| CW Doppler AV from Apical Views | 1,282 | 0.914 (0.914, 0.918) | 0.966 (0.966, 0.970) | 1.000 (1.000, 1.000) | 0.939 (0.930, 0.949) | |
| CW Doppler AV from the right parasternal view | 38 | 0.864 (0.861, 0.977) | 1.000 (0.995, 1.000) | 1.000 (1.000, 1.000) | 0.928 (0.874, 0.975) | |
| other | 74,885 | 0.999 (0.999, 0.999) | 0.998 (0.998, 0.998) | 0.986 (0.986, 0.987) | 0.998 (0.998, 0.998) | |

AV, aortic valve; CW, continuous wave Doppler; ITDS, internal test dataset; LVOT, left ventricle outflow tract; MV, mitral valve; PLAX, parasternal long-axis view. PSAX, parasternal short-axis view; PW, pulsed wave Doppler

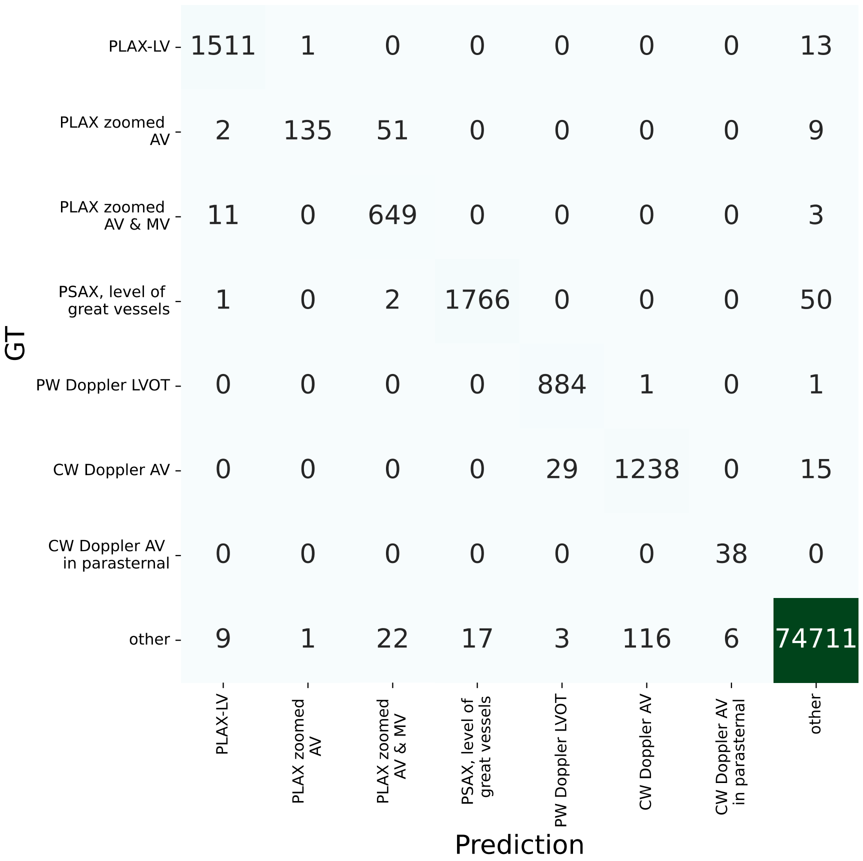
GT
PLAX-LV
PLAX zoomed AV
PLAX zoomed AV & MV
PSAX, level of great vessels
PW Doppler LVOT
CW Doppler AV
CW Doppler AV in parasternal
other
1511 1 0 0 0 0 0 13
2 135 51 0 0 0 0 9
11 0 649 0 0 0 0 3
1 0 2 1766 0 0 0 50
0 0 0 0 884 1 0 1
0 0 0 0 29 1238 0 15
0 0 0 0 0 0 38 0
9 1 22 17 3 116 6 74711
PLAX-LV
PLAX zoomed AV
PLAX zoomed AV & MV
PSAX, level of great vessels
PW Doppler LVOT
CW Doppler AV
CW Doppler AV in parasternal
other
Prediction

**DHDS**

| | n | Precision | Recall (Sensitivity) | specificity | F1-score | Accuracy |
|---|---|---|---|---|---|---|
| PLAX-LV | 3280 | 0.971 (0.971, 0.973) | 0.998 (0.998, 1.000) | 1.000 (1.000, 1.000) | 0.984 (0.981, 0.987) | |
| PLAX zoomed AV | 1313 | 0.989 (0.989, 0.994) | 0.909 (0.909, 0.914) | 0.999 (0.999, 0.999) | 0.947 (0.938, 0.956) | |
| PLAX zoomed AV & MV | 89 | 0.510 (0.509, 0.540) | 0.876 (0.875, 0.935) | 0.998 (0.998, 0.998) | 0.645 (0.589, 0.702) | |
| PSAX, level of great vessels | 3040 | 0.947 (0.947, 0.949) | 0.994 (0.994, 0.996) | 1.000 (1.000, 1.000) | 0.970 (0.966, 0.974) | |
| PW Doppler LVOT | 2182 | 0.999 (0.999, 1.000) | 0.998 (0.998, 1.000) | 1.000 (1.000, 1.000) | 0.998 (0.997, 1.000) | 0.995 (0.995, 0.995) |
| CW Doppler AV from Apical Views | 2275 | 0.987 (0.986, 0.989) | 0.968 (0.968, 0.970) | 0.999 (0.999, 0.999) | 0.977 (0.973, 0.982) | |
| CW Doppler AV from the right parasternal view | 20 | 0.810 (0.804, 1.000) | 0.850 (0.843, 1.000) | 1.000 (1.000, 1.000) | 0.832 (0.711, 0.930) | |
| other | 92460 | 0.998 (0.998, 0.998) | 0.997 (0.997, 0.997) | 0.987 (0.987, 0.987) | 0.997 (0.997, 0.998) | |

AV, aortic valve; CW, continuous wave Doppler; DHDS, Distinct Hospital Dataset; LVOT, left ventricle outflow tract; MV, mitral valve; PLAX, parasternal long-axis view. PSAX, parasternal short-axis view; PW, pulsed wave Doppler

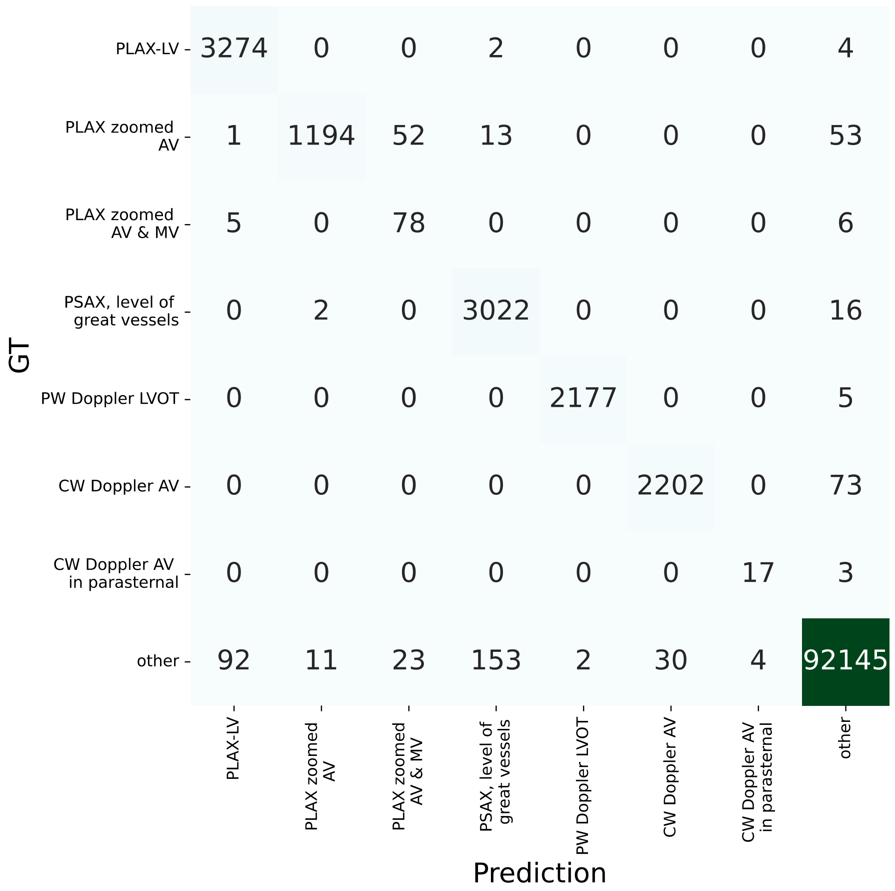

GT
Prediction
PLAX-LV
PLAX zoomed AV
PLAX zoomed AV & MV
PSAX, level of great vessels
PW Doppler LVOT
CW Doppler AV
CW Doppler AV in parasternal
other
3274 0 0 2 0 0 0 4
1 1194 52 13 0 0 0 53
5 0 78 0 0 0 0 6
0 2 0 3022 0 0 0 16
0 0 0 0 2177 0 0 5
0 0 0 0 0 2202 0 73
0 0 0 0 0 0 17 3
92 11 23 153 2 30 4 92145

**TDDS**

| | n | Precision | Recall (Sensitivity) | specificity | F1-score | Accuracy |
|---|---|---|---|---|---|---|
| PLAX-LV | 1377 | 0.984 (0.984, 0.988) | 0.992 (0.992, 0.996) | 1.000 (1.000, 1.000) | 0.988 (0.984, 0.992) | |
| PLAX zoomed AV | 193 | 0.907 (0.905, 0.968) | 0.404 (0.404, 0.428) | 0.998 (0.998, 0.998) | 0.560 (0.491, 0.625) | |
| PLAX zoomed AV & MV | 683 | 0.840 (0.840, 0.847) | 0.993 (0.992, 1.000) | 1.000 (1.000, 1.000) | 0.910 (0.900, 0.921) | |
| PSAX, level of great vessels | 1829 | 0.995 (0.995, 0.999) | 0.948 (0.948, 0.951) | 0.998 (0.998, 0.998) | 0.971 (0.965, 0.976) | |
| PW Doppler LVOT | 833 | 1.000 (1.000, 1.000) | 1.000 (1.000, 1.000) | 1.000 (1.000, 1.000) | 1.000 (1.000, 1.000) | 0.994 (0.994, 0.994) |
| CW Doppler AV from Apical Views | 1670 | 0.905 (0.905, 0.908) | 0.992 (0.992, 0.995) | 1.000 (1.000, 1.000) | 0.946 (0.939, 0.954) | |
| CW Doppler AV from the right parasternal view | 88 | 0.917 (0.915, 0.972) | 1.000 (0.998, 1.000) | 1.000 (1.000, 1.000) | 0.956 (0.926, 0.983) | |
| other | 67453 | 0.998 (0.998, 0.998) | 0.997 (0.997, 0.997) | 0.981 (0.981, 0.982) | 0.997 (0.997, 0.998) | |

AV, aortic valve; CW, continuous wave Doppler; TDDS, Temporally Distinct Dataset; LVOT, left ventricle outflow tract; MV, mitral valve; PLAX, parasternal long-axis view. PSAX, parasternal short-axis view; PW, pulsed wave Doppler

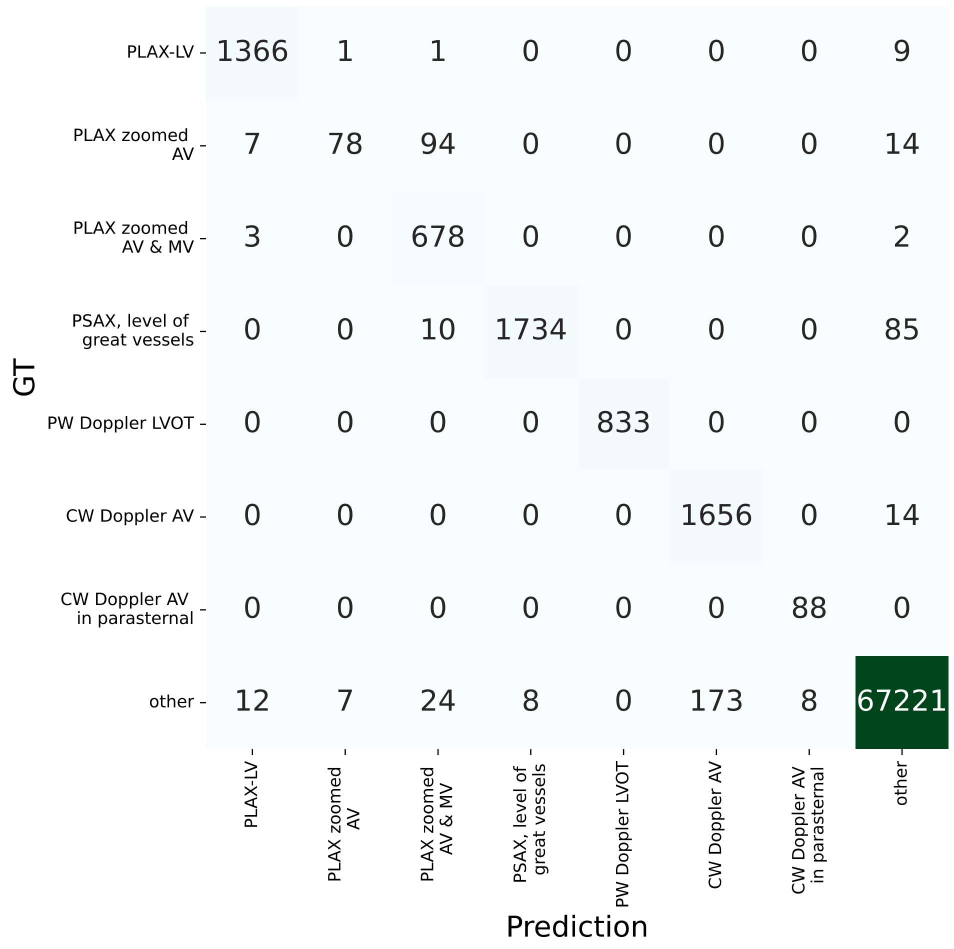
GT
Prediction
PLAX-LV
PLAX zoomed AV
PLAX zoomed AV & MV
PSAX, level of great vessels
PW Doppler LVOT
CW Doppler AV
CW Doppler AV in parasternal
other
1366 1 1 0 0 0 0 9
7 78 94 0 0 0 0 14
3 0 678 0 0 0 0 2
0 0 10 1734 0 0 0 85
0 0 0 0 833 0 0 0
0 0 0 0 0 1656 0 14
0 0 0 0 0 0 88 0
12 7 24 8 0 173 8 67221

**Supplemental Results 2. Distribution of DLi-ASc According to Conventional AS Parameters**

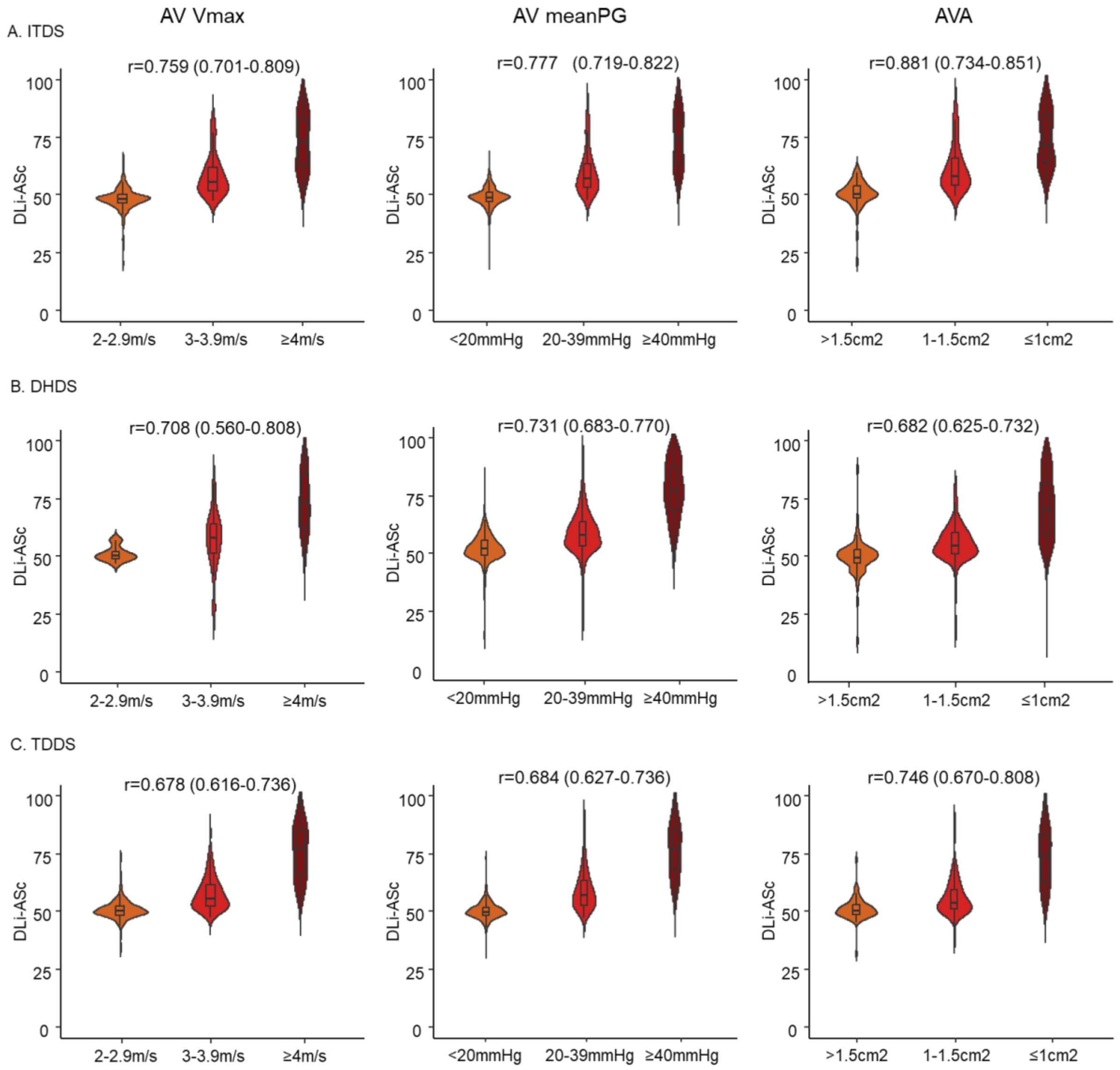

**Supplemental Results 3. Distribution of DLi-ASc in Discordant Cases of AS Severity**

In this study, discordant cases were defined as those where AV $V_{max}$, mPG, and AVA did not consistently fall into a single AS severity class, resulting in interpretations spanning two classes, such as mild to moderate, moderate to severe, or cases of low-flow, low-gradient AS where reduced stroke volume results in a lower pressure gradient despite significant stenosis. These cases were excluded from training, validation, and testing in the DDS. However, when included in the internal test dataset (ITDS) for comparison, the distribution of DLi-ASc was as follows. This result suggests that DLi-ASc can be helpful in cases where traditional parameters are discordant, making the assessment of AS severity challenging.

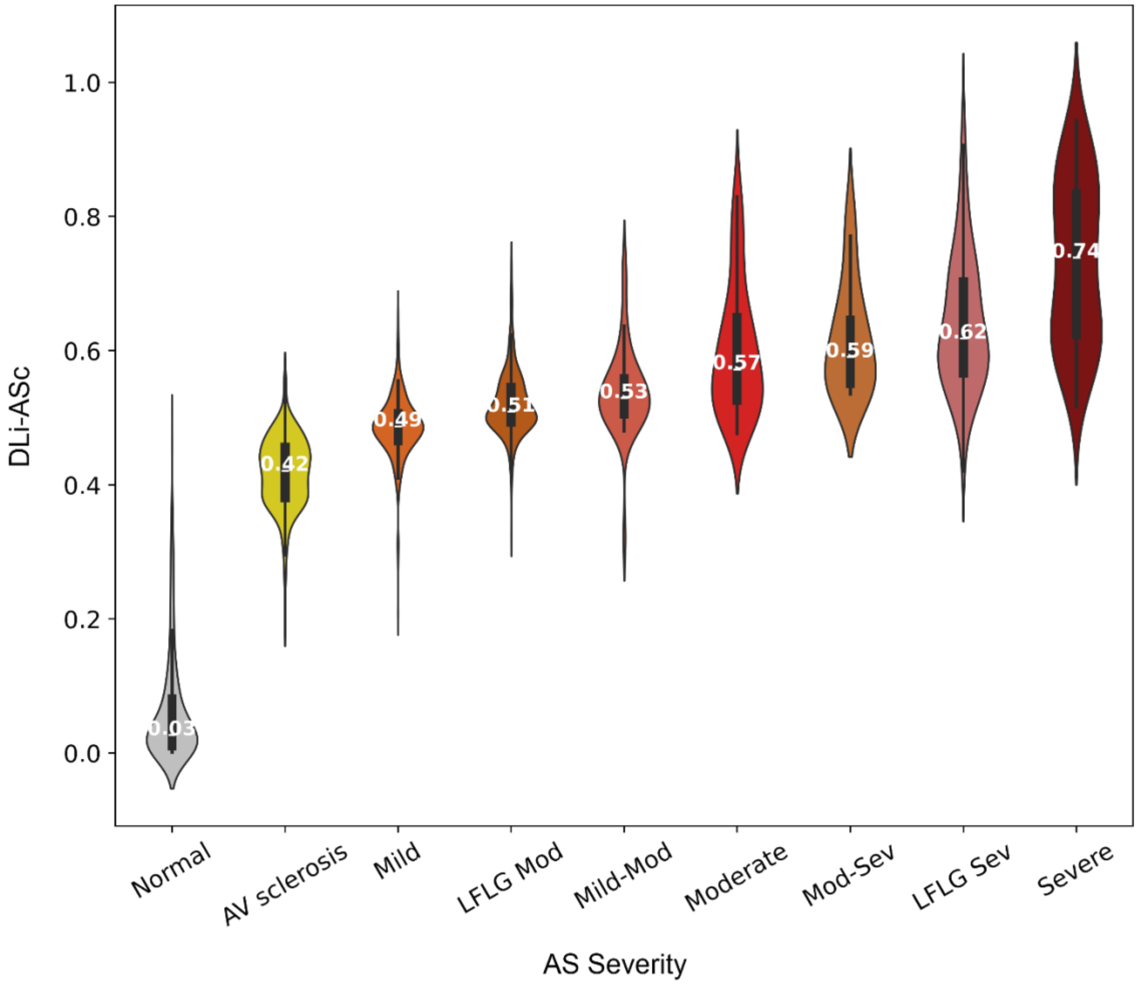


LFLG Mod, low-flow low-gradient moderate aortic stenosis; LFLG Sev, low-flow low-gradient severe aortic stenosis

**Supplemental Results 4. Discrimination of Low Flow Low Gradient Severe AS using DLi-ASc**

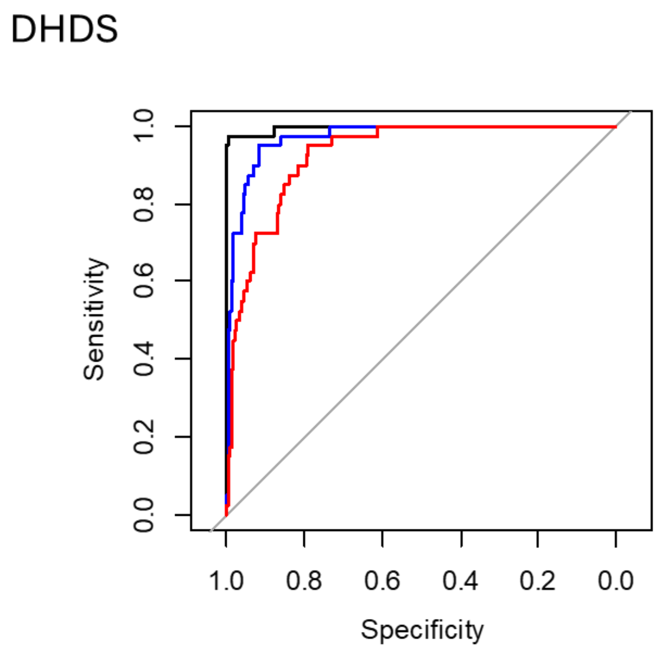


| | AUC (95% CI) |
|---|---|
| **Normal AV vs. LFLG severe AS** | 0.997 (0.991-0.999) |
| **Normal to mild AV vs. LFLG severe AS** | 0.974 (0.958-0.989) |
| **Normal to mod AV vs. LFLG severe AS** | 0.930 (0.902-0.958) |

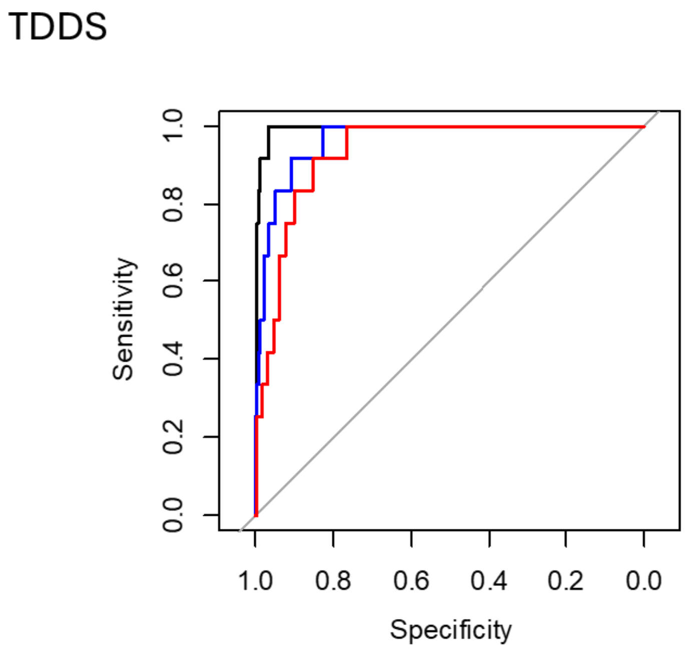


| | AUC (95% CI) |
|---|---|
| **Normal AV vs. LFLG severe AS** | 0.994 (0.987-0.999) |
| **Normal to mild AV vs. LFLG severe AS** | 0.966 (0.936-0.996) |
| **Normal to mod AV vs. LFLG severe AS** | 0.936 (0.895-0.976) |

**Supplemental Results 5. UMAP Visualization of AS Continuum Using Different Approaches**

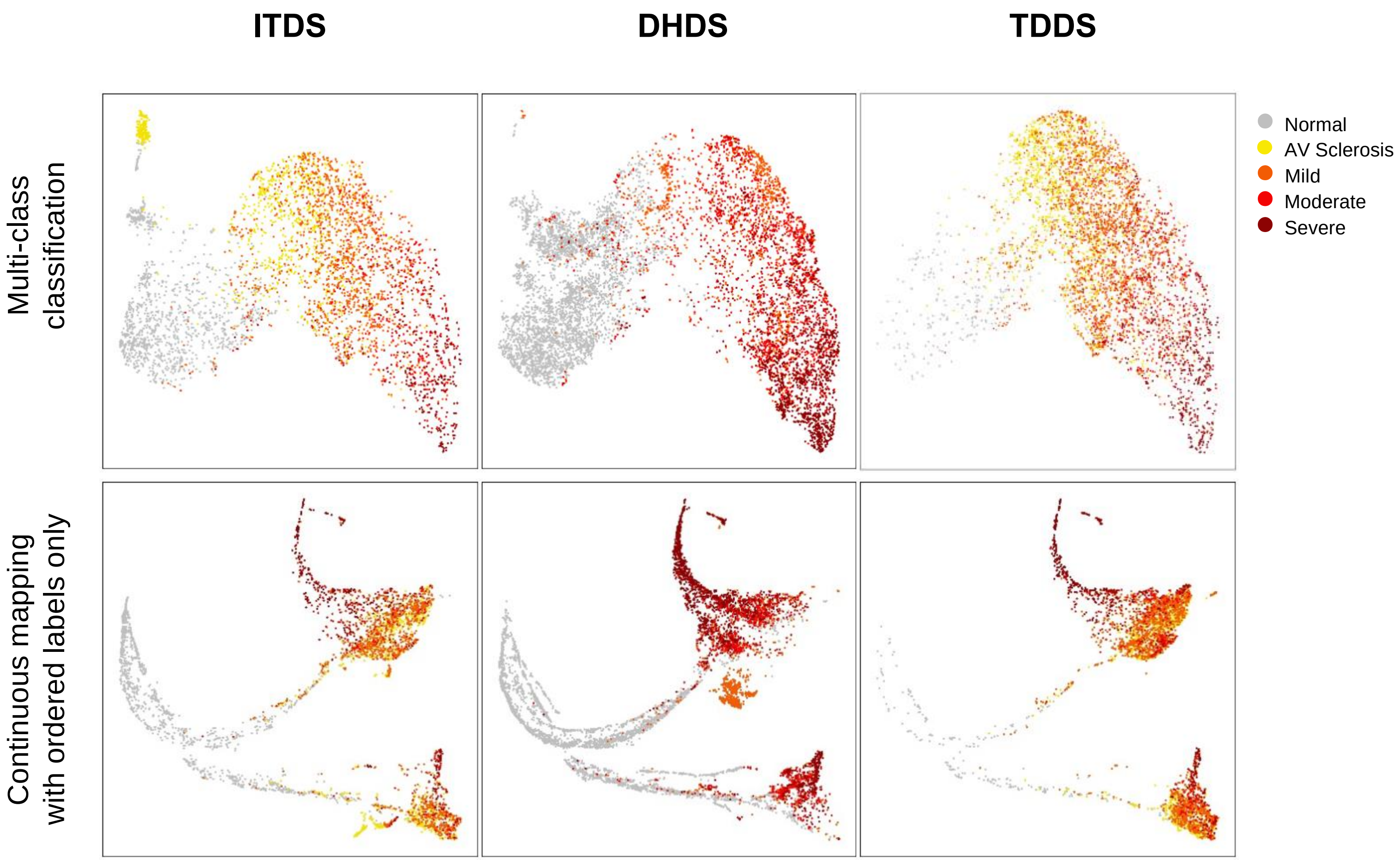


In the first row, multi-class classification uses standard 5-class cross-entropy loss with one-hot encoding. In the second row, the network is trained using only continuous mapping with ordered labels, without multi-task learning with auxiliary tasks.

**Supplemental Results 6. Representative Failure Cases**

The figure shows two representative cases where auto-measurement displayed significant discrepancies compared to manual measurement for AS parameters such as AV Vmax, mPG, and LVOT diameter.

In case A, measurement discrepancies in AV Vmax and mPG were attributed to ventricular bigeminy observed during AV Doppler measurement, leading to substantial beat-to-beat variability in the Doppler signal. For the manual measurement, AV Doppler was captured in a sinus rhythm, excluding premature beats, to ensure accuracy. In contrast, the auto-measurement processed all values in the AV CW Doppler signal and returned the highest value, resulting in the observed discrepancy. Despite this, both manual measurement and auto-measurement classified the case as severe AS. The estimated DLi-ASc Score was 87.2, further supporting the presence of severe AS.

In case B, there was a discrepancy in LVOT measurement due to differences in the measurement location. In manual evaluation, the measurement was taken slightly away from the aortic annulus toward the left ventricle, while the auto-measurement was performed closer to the annulus to determine the LVOT diameter. We previously evaluated various measurement points at different distances from the AV annulus in LVOT auto-measurement and found that measurement at the annulus level showed the highest correlation with the ground truth values (***Supplemental Methods*** *7*). However, in cases like this, where the LVOT demarcation is relatively unclear or the left ventricular septum is thick and sigmoid-shaped, the auto-measured LVOT value at the annulus level may be larger than usual. Nonetheless, both manual and automatic measurements classified the case as mild AS. The estimated DLi-ASc Score was 52.6, supporting the mild AS classification.

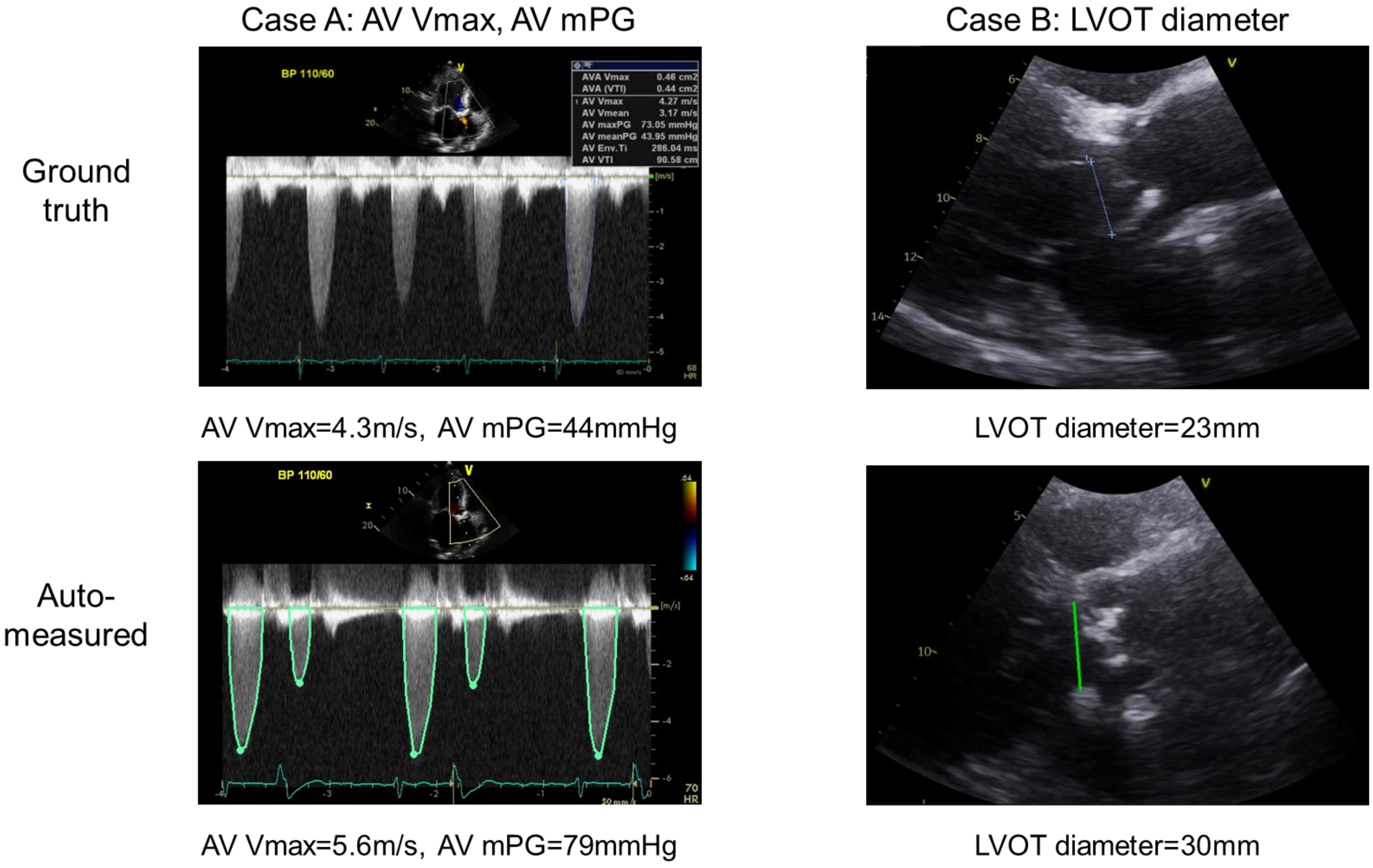
Case A: AV Vmax, AV mPG
Case B: LVOT diameter
Ground truth
Auto-measured
BP 110/60
AVA Vmax 0.46 cm2
AVA (VTI) 0.44 cm2
AV Vmax 4.27 m/s
AV Vmean 3.17 m/s
AV maxPG 73.05 mmHg
AV meanPG 43.95 mmHg
AV Env.Ti 286.04 ms
AV VTI 90.58 cm
AV Vmax=4.3m/s, AV mPG=44mmHg
LVOT diameter=23mm
AV Vmax=5.6m/s, AV mPG=79mmHg
LVOT diameter=30mm

**Supplemental Results 7. Comparative analysis with other existing studies**

We compared our AI algorithm with those from prior studies, specifically those by Holste et al. and Wessler et al. Key difference between our model and these earlier approaches are as follows.

- **Model Architecture and Data Processing:** Holste et al. utilized a 3D-ResNet 18 model with binary cross-entropy (BCE) loss for binary classification (non-severe vs. severe AS) on PLAX views, using 16 frames at a lower resolution (112 x 112). Additionally, Holste et al. used an ensemble of three models to improve robustness and classification accuracy. Wessler et al., in contrast, employed a WideResNet28 model for three-category classification (no AS, early AS, significant AS) using a single frame from PLAX and/or PSAX views. Our model employs an R2Plus1D18 backbone, sampling 16 frames at a higher resolution (224 x 224), with a continuum-aware multi-task loss. This approach allows our model to capture both spatial and temporal features effectively, facilitating more nuanced AS severity assessment.
- **Loss Function:** Unlike the previous studies that use binary or categorical cross-entropy loss, our model leverages a continuum-aware multi-task loss. This custom loss function not only accommodates the progressive nature of AS by mapping ordered severity labels, but it also includes auxiliary regression tasks to predict key TTE parameters. This dual approach enables a more comprehensive and clinically relevant assessment of AS severity.
- **Target Classification:** Our model's focus on a five-class continuum-based AS assessment provides finer granularity compared to the binary classification by Holste et al. or the three-class system of Wessler et al. By aligning better with clinical practices, our model supports a more precise and ordered understanding of AS severity, which is crucial for guiding treatment decisions.

| | Input type | Backbone | Loss | Required view | Sampled frame | Resolution | Target | Ensemble |
|---|---|---|---|---|---|---|---|---|
| Holste G et al. | Video | 3D-ResNet18 | CrossEntropyLoss | PLAX | 16 | 112 × 112 | non severe AS, severe AS | Yes, 3 models |
| Wessler BS et al. | Image | WideResNet28 | CrossEntropyLoss | PLAX or/and PSAX | 1 | 112 × 112 | no AS, early AS, significant AS | No |
| DLi-ASc (Ours) | Video | R2Plus1D18 | continuum-aware multi-task loss | PLAX or/and PSAX | 16 | 224 × 224 | Normal, AV sclerosis, Mild, Moderate, Severe | No |

The results shown below demonstrated the performance advantages of our approach across multiple datasets (ITDS, DHDS, and TDDS). Our model consistently achieved higher accuracy for detecting any AS, significant AS, and severe AS across all datasets, underscoring the effectiveness of our continuum-based methodology.

| | Holste G et al. | Wessler BS et al. | DLi-ASc (Ours) |
|---|---|---|---|
| ITDS | | | |
| any AS | 0.844 | 0.859 | 0.958 |
| significant AS | 0.947 | 0.784 | 0.979 |
| severe AS | 0.972 | - | 0.985 |
| DHDS | | | |
| any AS | 0.946 | 0.967 | 0.996 |
| significant AS | 0.934 | 0.832 | 0.969 |
| severe AS | 0.958 | - | 0.969 |
| TDDS | | | |
| any AS | 0.768 | 0.729 | 0.905 |
| significant AS | 0.914 | 0.714 | 0.949 |
| severe AS | 0.965 | - | 0.980 |

9. Loshchilov I, Hutter F. Sgdr: Stochastic gradient descent with warm restarts. *arXiv*

*preprint arXiv:160803983*. 2016.

10. Baumgartner HC, Hung JC-C, Bermejo J, et al. Recommendations on the echocardiographic assessment of aortic valve stenosis: a focused update from the European Association of Cardiovascular Imaging and the American Society of Echocardiography. *Eur Heart J Cardiovasc Imaging*. 2017;18:254-275.
